# Three Imaging Endophenotypes Characterize Neuroanatomical Heterogeneity of Autism Spectrum Disorder

**DOI:** 10.1101/2022.06.17.22276543

**Authors:** Gyujoon Hwang, Junhao Wen, Susan Sotardi, Edward S. Brodkin, Ganesh B. Chand, Dominic B. Dwyer, Guray Erus, Jimit Doshi, Pankhuri Singhal, Dhivya Srinivasan, Erdem Varol, Aristeidis Sotiras, Paola Dazzan, Rene S. Kahn, Hugo G. Schnack, Marcus V. Zanetti, Eva Meisenzahl, Geraldo F. Busatto, Benedicto Crespo-Facorro, Christos Pantelis, Stephen J. Wood, Chuanjun Zhuo, Russell T. Shinohara, Haochang Shou, Yong Fan, Nikolaos Koutsouleris, Raquel E. Gur, Ruben C. Gur, Theodore D. Satterthwaite, Daniel H. Wolf, Christos Davatzikos

**Author notes:** **Corresponding authors:** Gyujoon Hwang, PhD, Christos Davatzikos, PhD, 3700 Hamilton Walk, Philadelphia, PA 19104, USA /. These authors contributed equally to this work (Gyujoon Hwang, Junhao Wen, Susan Sotardi).

## Abstract

Autism spectrum disorder (ASD) is associated with high structural heterogeneity in magnetic resonance imaging (MRI). This work uncovers three neuroanatomical dimensions of ASD (*N*=307) using machine learning methods and constructs their characteristic MRI signatures. The presence of these signatures, along with their clinical profiles and genetic architectures, are investigated in the general population. High expression of the first dimension (A1, “aging-related”) is associated with globally reduced brain volume, cognitive dysfunction, and aging-related genetic variants. The second dimension (A2, “schizophrenia-like”) is characterized by enlarged subcortical volume, antipsychotic medication use, and partially overlapping genetic underpinnings to schizophrenia. The third dimension (A3, “classical ASD”) is distinguished by enlarged cortical volume, high non-verbal cognitive performance, and genes and biological pathways implicating brain development and abnormal apoptosis. Thus, we propose a three-dimensional endophenotypic representation to construe the heterogeneity in ASD, which can support precision medicine and the discovery of the biological mechanisms of ASD.

## 1. Main

The current definition of autism spectrum disorder (ASD) encompasses a range of social communication deficits and restricted, repetitive behaviors.^1^ In the past few decades, its diagnostic criteria evolved from being narrow and rare to being more inclusive and common, based on a new understanding of the complex cognitive and behavioral traits found in affected individuals. Its global prevalence is estimated to be 1%,^2^ but this estimate varies greatly within and across nations and sociodemographic groups, which may be attributed to the lack of consensus on the historical definition of ASD and different levels of awareness of the diagnosis.

Autistic individuals exhibit considerable heterogeneity in clinical symptoms, brain neuroanatomy and function, and genetics. The broad diagnostic criteria for ASD result in a vast array of clinical presentations with respect to intelligence and communication abilities, as well as its severity and co-occurring disorders.^3^ Prior neuroimaging research has also demonstrated significant structural heterogeneity within individuals with ASD compared to typically-developing (TD) individuals.^4^ Overall, enlarged total brain volume has been primarily observed in children with ASD from case-control analyses, followed by a slow growth into adolescence and adulthood.^5^ However, these findings diverge at a localized level^6^ with significant inter-individual variability.^7^ Additionally, genetic heterogeneity has been evident in ASD.^8^ Although more than 100 genes have been associated with ASD, the extent to which these loci collectively or independently contribute to the neuroanatomical heterogeneity, under distinct mechanisms and pathways, is not yet fully understood.^9^ Therefore, treating ASD as one uniformly-defined disorder hinders precision medicine.

Individuals with ASD are also at risk of developing co-occurring psychiatric conditions that further introduce heterogeneity, including major depressive disorder (MDD), schizophrenia (SCZ), bipolar disorder, obsessive-compulsive disorder (OCD), attention-deficit/hyperactivity disorder (ADHD), and myriad physical health conditions, including diabetes mellitus (DM), hypertension, gastrointestinal and pulmonary disorders.^10^ Among these conditions, the rates of SCZ and DM increase significantly with age.^11^ As a result, a wide variety of medications are prescribed to autistic individuals to manage comorbidities, while no specific pharmacological treatment exists for ASD itself.^12^ Many of these increased comorbid risks are persistent even after controlling for demographic and lifestyle-related factors,^13^ which suggests that they may share some biological processes with ASD. For example, ASD and many other psychiatric disorders share partially overlapping genetic risk factors.^14^

Clustering seeks to identify distinct patterns within a population. Numerous clustering methods, mostly traditional unsupervised methods, have been applied to dissect neuroanatomical heterogeneity in ASD.^15,16^ One critical limitation is that these traditional models perform clustering in the case population alone, thereby ignoring the specific ways in which autistic individuals vary with respect to the norm. In contrast, semi-supervised clustering methods^17-19^ seek the “1-to-*k*” mapping (*k* is the number of clusters) from the domain of a reference group (i.e., TD) to the atypical group (i.e., ASD), thereby clustering the atypical group according to the deviating patterns and alleviating confounds unrelated to the atypicality. These methods have been successfully applied to Alzheimer’s Disease,^19^ late-life depression,^20^ and SCZ,^21^ but are currently scarce in the ASD literature.^22,23^ In addition, semi-supervised methods dissect this heterogeneity into a multidimensional representation on an individual basis, whereby a continuous “dimension score” measures the degree to which patterns deviate from the norm along the given dimension.

The current study aims to characterize heterogeneity in neuroimaging and clinical profiles among late adolescents to adults with ASD (ages between 16 and 64), condense this heterogeneity into an endophenotypic, multidimensional representation, and investigate the expression of the endophenotypes in the general population. First, the semi-supervised HYDRA (Heterogeneity through Discriminative Analysis) model^17^ was trained using T1-weighted magnetic resonance images (MRI) of 307 individuals with ASD and 362 TD controls from the ABIDE (Autism Brain Imaging Data Exchange) consortium^24^ and the *k* dimension scores were derived for each dimension and individual. The HYDRA model was then applied to participants from the PHENOM consortium^21^ and the UK Biobank^25^ to investigate the expression of these ASD neuroanatomical dimension scores in both individuals with a diagnosis of SCZ and the general population. We hypothesized that 1) multiple distinct dimensions coexist to account for the underlying neuroanatomical, clinical, and genetic heterogeneity in ASD, and 2) the dimension scores which may serve as imaging-derived endophenotypes are potentially prominent also in non-ASD populations.

## 2. Results

### 2.1. The neuroanatomy of ASD is heterogeneous

Using T1-weighted MRI data from the ABIDE sample (*N*=669) (***Table 1***), we examined the neuroanatomical heterogeneity in ASD, via group-level comparisons of 145 regions-of-interest (ROIs) (***Supplementary Table 1***) between the ASD and TD groups, controlling for age, sex, and the imaging site. Group-wise differences were more prominent for the variance of the brain volumes than for the mean (***Supplementary Fig. 1***). The left lateral ventricle showed the most significant mean volume increase in ASD compared to TD (Cohen’s *d*=0.32, *P*_*FDR*_=0.001, *t*-test without assuming equal variance), followed by the 3^rd^ ventricle, left planum temporale, and left planum polare (*d*=0.23, *P*_*FDR*_<0.02), all with significantly increased variance (*P*_*FDR*_<1×10^−5^, *F*-test for equal variances).

**Table 1.**
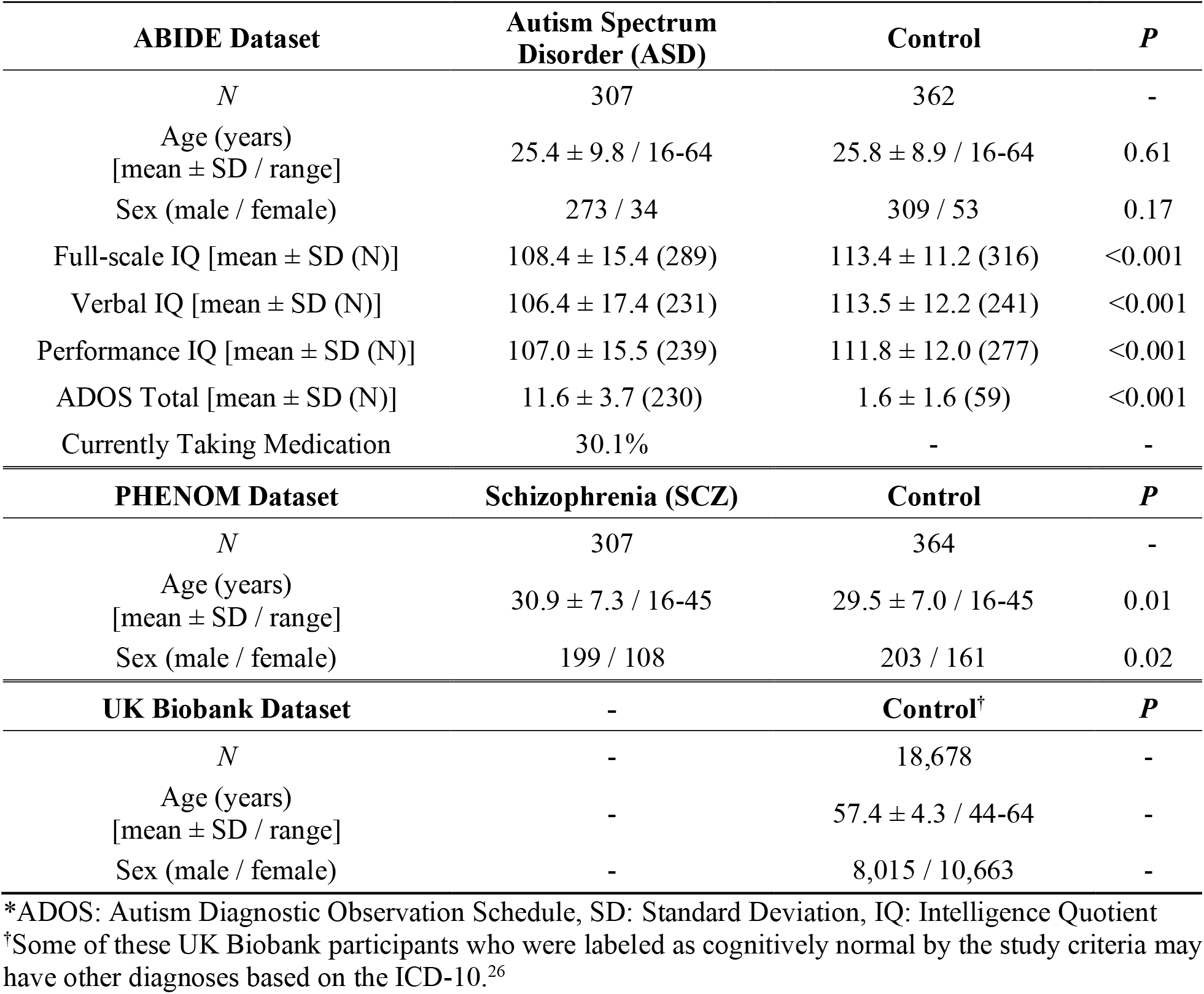
Demographic and clinical summary of the datasets.

#### 2.1.1. Search for a reproducible HYDRA model

HYDRA was trained with the 145 ROIs of the ABIDE participants to define the *k* neuroanatomical dimensions of ASD. The three-dimensional (*k*=3) scheme was optimal to best capture the heterogeneity, supported by four reproducibility analyses. First, this dimensional scheme showed a relatively high adjusted Rand index (ARI) (0.52±0.04).^27^ Second, this index was the most significantly different from the null distribution of ARIs computed by permuting cases and controls (*P*_*FDR*_<1E-8, 100 runs) (***Fig. 1a***). Third, the three dimension scores computed by the HYDRA model exhibited relatively superior reproducibility between the cross-validated (CV) runs (***Fig. 1b***). Finally, the split-sample validation showed high correspondence (***Supplementary Fig. 2***). Therefore, results from the three-dimensional HYDRA model will be discussed hereafter, with A1, A2, and A3 denoting the three-dimensional representation (***Fig. 1c*** and ***Supplementary Video 1***).

**Fig. 1:**
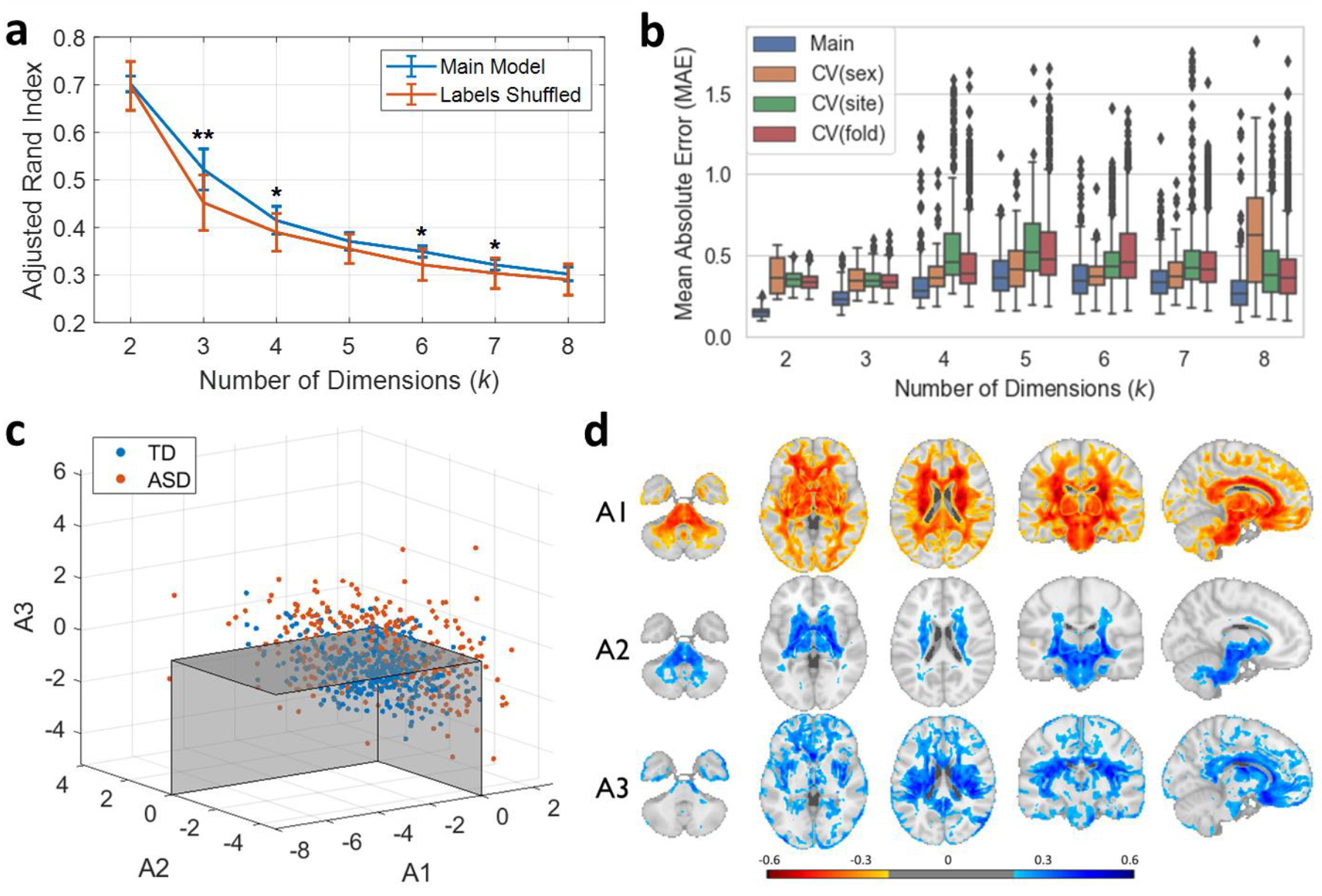
Three reproducible neuroanatomical dimensions of ASD. (a) The three-dimensional (*k*=3) clustering scheme showed a relatively high adjusted Rand index (ARI) and exhibited the most significant permutation test results (100 runs). (b) Mean absolute error (MAE) between pairwise ASD dimension scores computed from different random initializations including all subjects (blue bars, 50 runs each) and computed from different cross-validation (CV) folds (20 runs per fold) are plotted. Note that as the dimensions are aligned and labeled based on the pairwise MAE, a better average MAE is expected at random for a higher number of dimensions. (c) The three-dimensional representation of the neuroanatomical heterogeneity in ASD: both TD (*N*=362) and ASD (*N*=307) brains are projected onto the final three-dimensional space. The boundaries are drawn with the threshold of zero for visualization. (d) The color maps are based on Pearson correlations between dimension scores and voxel-wise regional volumetric (RAVENS) statistics (gray and white matters combined, without the cerebrospinal fluid) using all subjects (*N*=669) masked by *P*_*Bonferroni*_<0.05. \**P*_*FDR*_<1E-3 \*\**P*_*FDR*_<1E-8

#### 2.1.2. Three neuroanatomical dimensions of ASD are revealed

Neuroanatomical characteristics of the three ASD dimensions were evaluated to gain post hoc intuition about the most pertinent imaging patterns that constitute each dimension (***Fig. 1d***). A high A1 dimension score was associated with a widespread pattern of smaller volumes except for the orbital part of the inferior frontal gyrus (pars orbitalis). The volume loss in the white matter and the subcortex was more pronounced than the gray matter. A high A2 dimension score was associated with larger subcortical structures, especially the pallidum and the internal capsule. A high A3 dimension score was associated with a widespread pattern of larger volumes, particularly involving the frontal gray matter and insula. The A1 and A3 dimensions both involved proportionally larger gray matter, while being characterized by small and large total brain sizes respectively (***Supplementary Fig. 3***).

#### 2.1.3. Three neuroanatomical dimensions show distinct clinical profiles

The age of individuals with ASD was positively correlated with their A1 dimension score at uncorrected levels (*r*=0.13, *N*=307, *P*=0.026). Note that age and sex were regressed out from the ROIs before training using association parameters estimated in the TD controls. Performance IQ (or the non-verbal IQ; Intelligence Quotient)^28^ was inversely correlated with the A1 score at uncorrected levels (*N*=239, *r*=-0.136, *P*_*FDR*_=0.085) and positively with the A3 score (*r*=0.124, *P*_*FDR*_=0.085) (***Fig. 2a*** and ***Supplementary Table 2***). Variations of the administered IQ tests are listed in ***Supplementary Table 3***. The ADOS (Autism Diagnostic Observation Schedule)^29^ total score was positively correlated with the A3 score at uncorrected levels (*N*=230, *r*=0.142, *P*_*FDR*_=0.095). Individuals with ASD taking antipsychotics (*N*=15) received significantly higher A2 scores than the others (*N*=188, *P*_*FDR*_=0.048, Cohen’s *d*=0.65). A similar trend was observed for those taking medications for OCD (*N*=18, *P*_*FDR*_=0.095, *d*=0.53) (***Supplementary Table 4***). Except for the ADHD medications (*P*_*FDR*_=0.02, two-sample *t*-test; younger individuals took them at a higher rate), age was not significantly related to the medication status in any medication type (*P*>0.05).

**Fig. 2:**
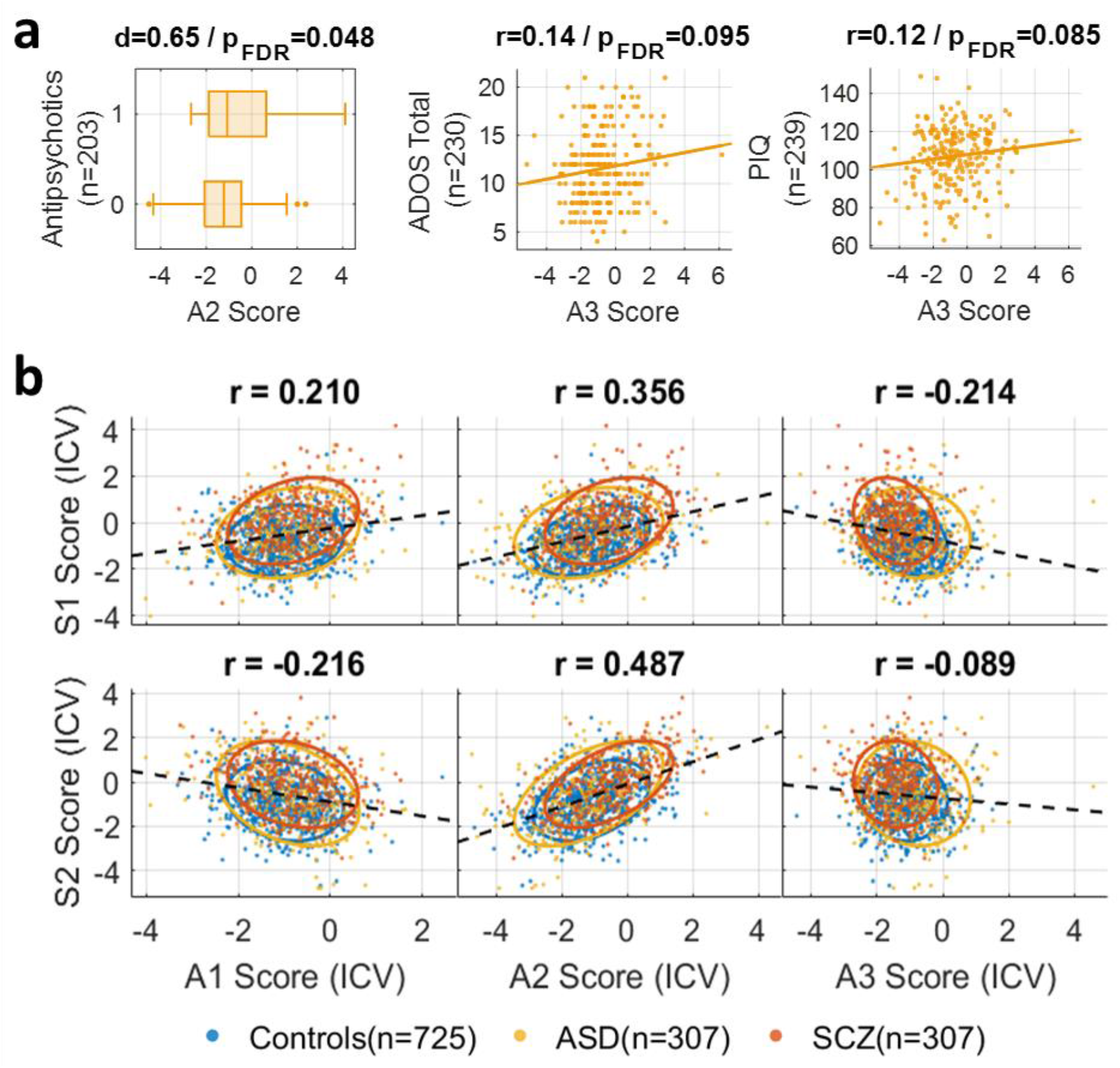
Association between the A2 dimension and schizophrenia (SCZ). (a) Among individuals with ASD, a higher A2 score was associated with taking antipsychotics, while a higher A3 score was associated with both the ADOS (Autism Diagnostic Observation Schedule) total score and the performance IQ. (b) Correlations between the ASD and SCZ dimension scores are plotted by diagnosis using the combined dataset of ABIDE and PHENOM, after the scores were adjusted for the intracranial volume (ICV).

#### 2.1.4. The ASD dimensions are correlated with SCZ dimensions

To investigate the imaging-derived dimensional correlations between ASD and SCZ, we applied i) the trained HYDRA model on ABIDE to the PHENOM participants and ii) the trained HYDRA model on PHENOM (from Ganesh et al.^21^; *k*=2) to the ABIDE participants. The latter derived expressions of the two SCZ dimension scores (S1 and S2) for each participant. In our previous work, the S1 dimension was characterized by smaller brain volumes, especially in the frontal, temporal and insular cortices, while the S2 dimension was characterized by larger basal ganglia and internal capsule.^21^ ***Supplementary Fig. 4*** shows expressions of these two SCZ dimensions in individuals with ASD.

The A1 dimension score was positively correlated with the S1 dimension score (*r*=0.210, *N*=1,339, *P*_*FDR*_<1×10^−8^; ICV-corrected), although the A1 entailed relatively smaller white matter and subcortex than the S1 (***Fig. 2b, Supplementary Fig. 4 and 5***). The A2 dimension score was positively correlated with both S1 and S2 dimension scores (*P*_*FDR*_<1×10^−8^; ICV-corrected), though more strongly with the S2. Both the A2 and S2 dimensions demonstrated larger subcortical structures, especially the bilateral pallidum regions.

### 2.2. Expression of the three ASD dimensions in the general population

To examine the three neuroanatomical dimensions of ASD in the general population, we applied the trained HYDRA model on ABIDE to the UK Biobank participants (*N*=18,678)^25^ and derived the expressions of the three dimensions. The score distributions were comparable to the distributions of the control groups of ABIDE and PHENOM (***Supplementary Fig. 6***).

#### 2.2.1. The clinical profiles of the three ASD dimensions

Among the UK Biobank participants, the A1 dimension score was significantly correlated with the brain age gap (predicted age based on the structural brain minus the chronological age,^30^ *N*=10,894, *r*=0.31, *P*_*FDR*_<1×10^−8^ with both raw and ICV-corrected A1 score) (***Supplementary Table 5***). Individuals with high A3 scores performed significantly better on four neuropsychological tests that were examined compared to those with low scores, especially the fluid intelligence (*N*=17,684, *r*=0.14) and Digit Span Forward (*N*=12,294, *r*=0.12),^31^ while the opposite trends were found with the A1 scores (all *P*_*FDR*_<1×10^−8^). Those with diabetes mellitus (DM, *N*=742, 4%) received significantly higher A1 scores compared to those without (Cohen’s *d*=0.34, *P*_*FDR*_<1×10^−8^; age- and sex-matched) (***Fig. 3***). ***Supplementary Table 2*** provides the complete table for the clinical associations.

**Fig. 3:**
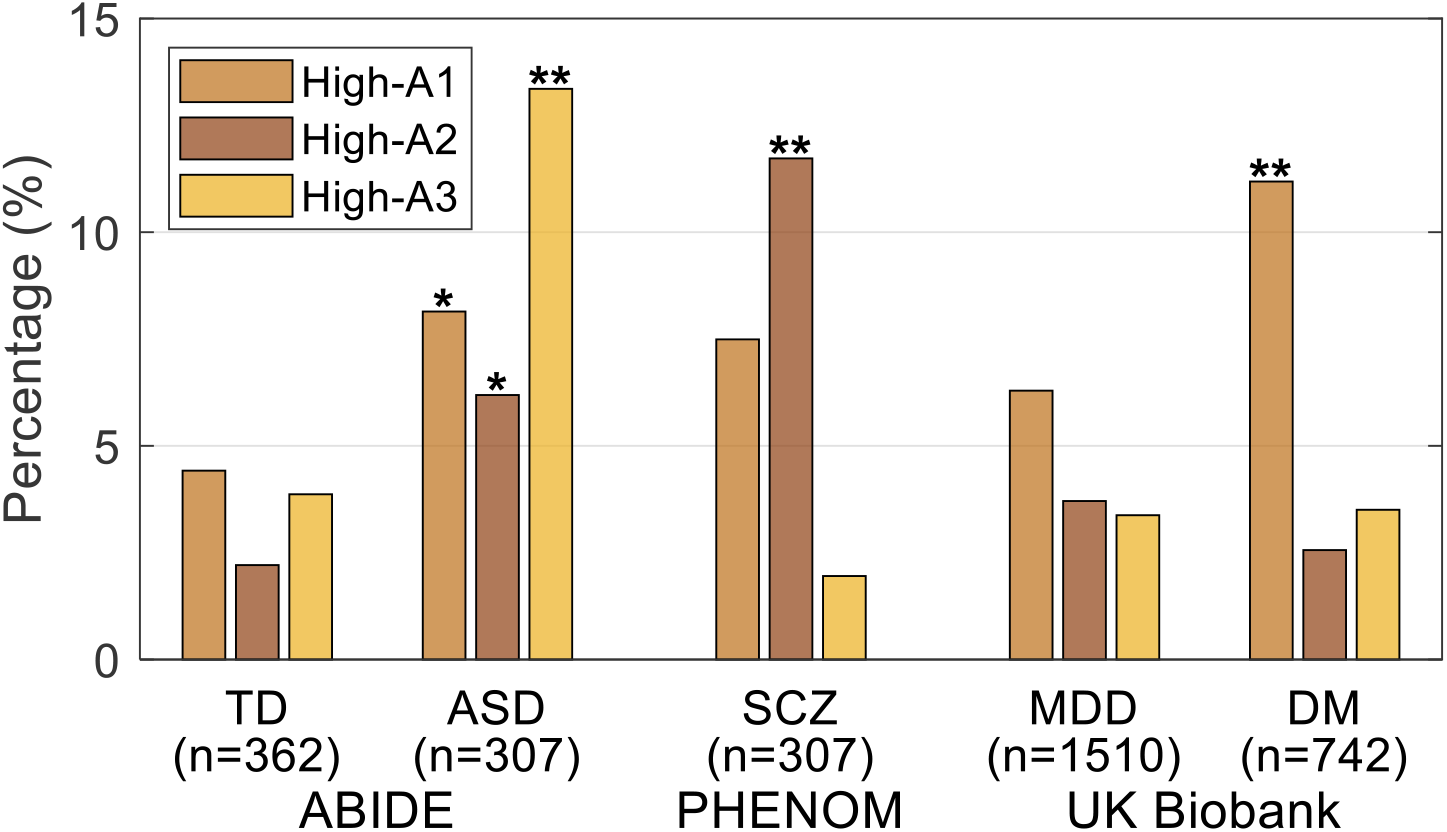
Association between the A1 dimension and diabetes in the general population. ASD dimension scores in different diagnostic groups are summarized: TD and ASD groups from ABIDE, SCZ from PHENOM, and MDD (Major Depressive Disorder) and DM (Diabetes Mellitus) from the UK Biobank. Each diagnosis group was segmented into three subgroups using a score threshold (=0.96). For example, the High-A1 subgroup included individuals with A1>A2 and A1>A3, while A1>threshold. The threshold was set at where the sum of the three bars of the TD group would equal 10%. * *P*_*FDR*_<0.05/ ** *P*_*FDR*_<0.001 (Chi-squared tests based on the rate of each bar compared to TD)

#### 2.2.2. Differences in genome-wide associations

The A1 dimension was significantly associated with one novel lead SNP (rs10706986; single nucleotide polymorphism); the A2 dimension, two lead SNPs (rs1062034, novel; rs1160248); the A3 dimension, one lead SNP (rs72810098) (***Table 2*** and ***Fig. 4a-c***). The candidate SNPs in the two previously identified genomic loci were reported to be associated with a wide range of clinical traits, including brain volumetric measures (A1 and A2), schizophrenia (A2), cognitive performance (A2), and sex hormone-binding globulin levels (A3) in the GWAS Catalog^32^ (***Supplementary Files 1-3***). The GWAS results for the schizophrenia S1 and S2 dimensions are also presented for comparison in ***Supplementary Fig. 7***. Quantile-quantile plots are presented in ***Supplementary Fig. 8***. The definitions of the lead, candidate, and independent significant SNPs, and genomic risk loci are presented in ***Supplementary Notes 1***.

**Table 2.** Genomic risk loci for the three ASD dimensions in the general population (UK Biobank)

| ASD Dimension | Lead SNP | $P$ | Minor Allele | $\beta$ | Previously reported |
| --- | --- | --- | --- | --- | --- |
| A1 | rs10706986 | $2.63 \times 10^{-11}$ | T | -0.08 | No |
| A2 | rs1062034 | $3.29 \times 10^{-9}$ | G | -0.08 | No |
| | rs1160248 | $3.09 \times 10^{-8}$ | C | 0.06 | Yes |
| A3 | rs72810098 | $3.10 \times 10^{-8}$ | C | -0.19 | Yes |

**Fig. 4:**
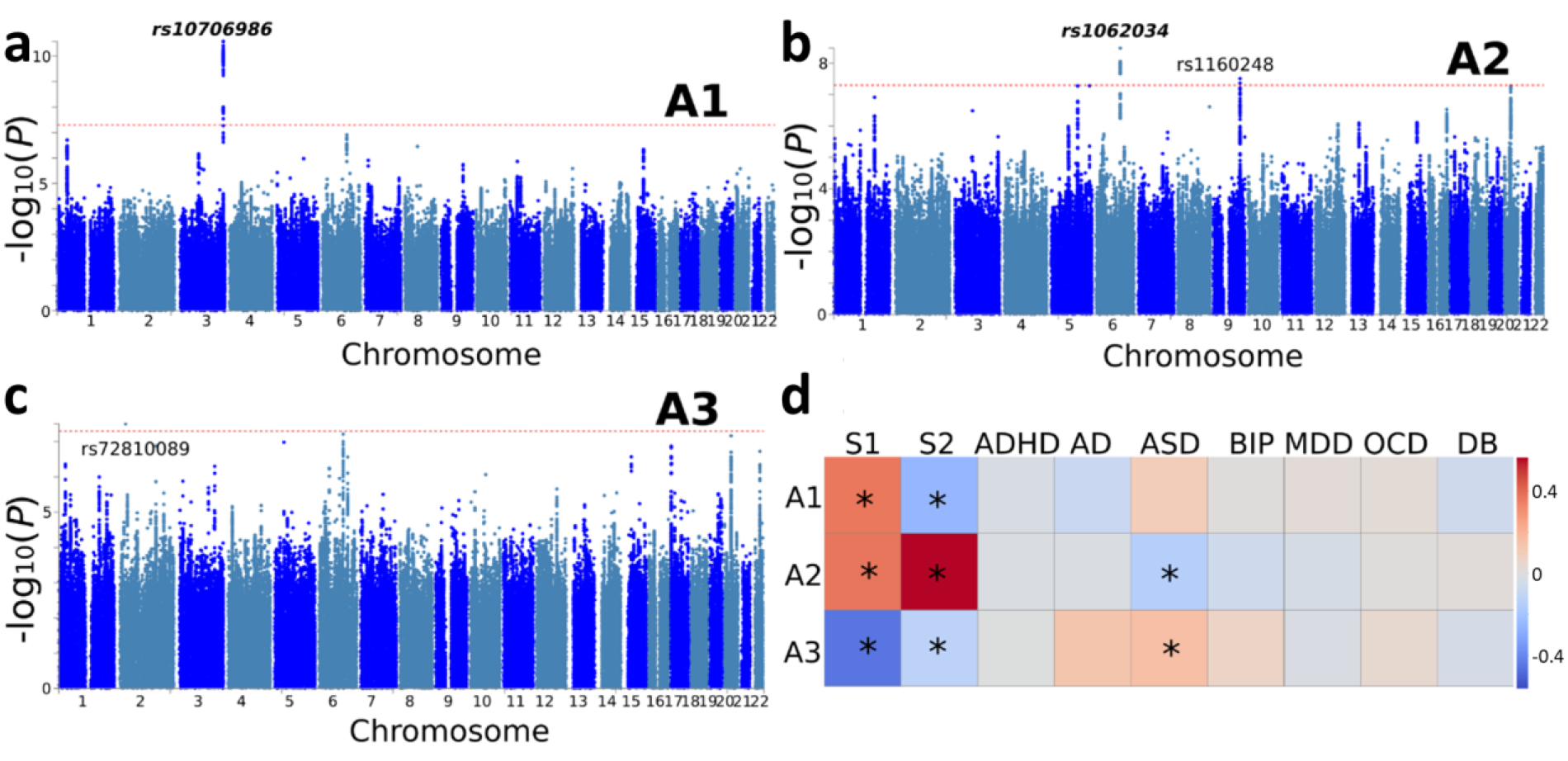
Genetic architecture of the three ASD dimensions in the general population. We refer to a lead SNP as novel (bold and italic) if it has not been associated with any clinical traits in the GWAS Catalog. The genome-wide default *P*-value threshold (*P*<5×10^−8^, dotted red lines) was used for all GWAS. (a) Genomic loci/lead SNP of the A1 dimension. (b) Genomic loci/lead SNPs of the A2 dimension. (c) Genomic loci/lead SNP of the A3 dimension. (d) Genetic correlations between ASD dimensions, SCZ dimensions, and other brain disorders. Genetic correlations were estimated using LDSC^35^ (unbiased from population overlaps). In particular, the GWAS summary statistics of ASD and SCZ dimensions were derived from the current study, and those of the other disease traits were from previous GWAS studies (see elsewhere^36^ for details of the inclusion criteria.) \**P*<0.05 in the genetic correlation estimates. ADHD: attention-deficit/hyperactivity disorder; AD: Alzheimer’s Disease; BIP: bipolar disorder; MDD: major depressive disorder; DB: diabetes mellitus.

For gene-level associations, the A1 dimension was significantly associated with the *CDC20, MPL, GMNC, HYI*, and *FOXO3* genes; the A2 dimension, the *DEFB124, FOXO3, REM1, PAPPA, DEFB119, STX6*, and *SELO* genes; the A3 dimension, the *C20orf112* and *METTL16* genes (gene-level *P*<2.64×10^−6^) (***Supplementary Table 6***). Gene-level Manhattan plots are presented in ***Supplementary Fig. 9***.

From the gene-set enrichment analysis (a hypothesis-free approach),^33^ the A2 dimension was enriched in the biological pathway of hydrogen peroxide-mediated programmed cell death (gene-set-level *P*=3.78×10^−6^, *β*=2.86); the A3 dimension, the biological pathway regulating the intrinsic apoptotic signaling pathway in response to DNA damage by p53 class mediator (*P*=4.22×10^−6^, *β*=0.83). For the A2 dimension, the expression levels were significantly enriched in many tissue types (a prioritized gene approach),^34^ including brain (e.g., cerebellum) and tibial nerves, for the following genes: *FOXO3* and *SESN1* ***(Supplementary Fig. 10)***.

#### 2.2.3. ASD dimensions are highly heritable in the general population

The three dimension scores of ASD showed highly significant SNP-based heritability estimates (*h*^*2*^): 0.38±0.04, 0.71±0.04, and 0.55±0.04, respectively (all *P*<10^−4^). For the SCZ dimensions, they were 0.40±0.04 and 0.51±0.04, respectively (*P*<10^−4^).

#### 2.2.4. Genetic covariance between ASD dimensions and other clinical traits

The A2 dimension showed positive genetic correlations with both S1 (*r*_*g*_=0.37, *P*=0.001) and S2 dimension (*r*_*g*_=0.56, *P*<10^−8^), and a negative genetic correlation with ASD (*r*_*g*_=-0.15, *P*<0.01). The A3 dimension showed a positive genetic correlation with ASD (*r*_*g*_=0.16, *P*<0.02) (***Fig. 4d***).

## 3. Discussion

We propose a three-dimensional endophenotypic representation to distill the neuroanatomical and clinical heterogeneity in the ASD population. Furthermore, we have investigated the expression of the three ASD dimensions in a population with a diagnosis of SCZ and the general population. Our results substantiate considerable neuroanatomical and clinical heterogeneity across the three ASD dimensions, their partially overlapping imaging patterns and genetic underpinnings compared to the two SCZ dimensions, and their heterogeneous expressions in the general population.

### 3.1. Three-dimensional representation of ASD neuroanatomical heterogeneity

We discovered three machine learning-derived, reproducible neuroanatomical dimensions of ASD. The dimension most significantly associated with cognitive impairment (A1) was characterized by smaller brain size except for the orbital part of the inferior frontal gyrus (pars orbitalis). The inferior frontal gyrus is a primary language processing region that has been extensively studied in ASD^37^ and the differential role of the pars orbitalis compared to the rest of the inferior frontal gyrus in ASD has been reported from case-control studies.^38^ The A1 dimension may facilitate the future investigation of this abnormality. While both the A1 and S1 dimensions showed a widespread brain volume loss, the atrophy was more severe in the subcortex and the white matter for the A1 dimension. This difference in the gray-to-white matter volume ratio was previously observed in comparisons of ASD and SCZ.^39^

The second dimension (A2) showed enlarged subcortical volumes, which resembled a previously reported S2 neuroanatomical dimension of SCZ.^21^ ASD as combined group has been associated with smaller subcortical volumes,^6^ and thus, examining the individuals with high A2 scores separately may be useful, potentially revealing transdiagnostic characteristics between ASD and SCZ. Overlapping regions of structural alteration are commonly seen between many neuropsychiatric disorders, including ASD and SCZ at the group level.^40^ However, these analyses were most likely complicated by the underlying heterogeneity in the ASD neuroanatomy. A relatively large proportion of affected individuals with a given neuroanatomical signature may dominate an analysis when averaged across a group exhibiting different structural patterns.

The third dimension (A3) was characterized by larger cortical volume, especially the frontal gray matter and insula. Increased frontal gray matter is consistent with reports from many case-control studies of ASD,^5^ while the increased insular volume has been less evident.^6^ However, numerous studies report atypical activation and connectivity of the insular cortices,^41^ and therefore, future studies may benefit from examining the individuals with high A3 scores separately. In the context of the developmental trajectory of ASD brains,^5^ the A3 dimension may be interpreted as the degree of resilience to the impaired growth that follows the initial overgrowth.

Detailed comparison of these three dimensions to the few previously identified neuroanatomical dimensions or clusters of ASD is premature.^15,16^ This is, in part, due to the inconsistencies regarding the age ranges of the sample, imaging modalities, and clustering techniques. Despite these discrepancies, some convergence in findings has been noted.^16^ First, most sets of neuroimaging dimensions of ASD indicate a combination of both increases and decreases in structural and functional neuroimaging signatures, instead of pointing in a uniform direction. Second, most dimensions are characterized by spatially distributed imaging patterns, instead of isolated or focal patterns. Our findings corroborate the significant neuroanatomical heterogeneity in individuals with ASD, distilling such heterogeneity into a three-dimensional representation. This representation quantifies disease susceptibility scores at the individual level – a prototype for precision medicine if extensively validated – and provides an instrument to elucidate distinct mechanisms of ASD.

### 3.2. Clinical profiles of the three dimensions

One major challenge of building a heterogeneity-aware multidimensional representation of a clinical population is the lack of ground truth — all clustering methods lead to a solution. Thus, cross-validation, replications using unseen data, and cross-domain validations (e.g., defining the dimensions in neuroimaging and applying them to clinical or genomic variables) are indispensable for any claims.^42^ The final dimensions must ideally be meaningful by adding value to understanding the symptoms or the etiology. To this end, we investigated the clinical profiles of the three ASD dimensions.

The expression of the A1 dimension was correlated with age and lower performance IQ. Similarly, in the general population, it was associated with cognitive impairment, as well as the brain age gap and the rate of DM, which have been linked to a wide range of age-related brain deteriorations.^43^ There are numerous reports of clusters based on clinical symptoms of ASD, and most indicate at least one cluster exhibiting lower cognitive performance or intelligence, which may correspond to the A1 dimension.^15^ ASD also carries many co-occurring psychiatric and physical health conditions.^10^ This study suggests that individuals with high A1 scores, both ASD and non-ASD alike, may be prone to developing some of these conditions, due to partially overlapping neurobiological processes.

The A2 dimension score was significantly correlated with the use of antipsychotics among individuals with ASD. Further, this dimension was significantly correlated with both the S1 and S2 dimensions derived from SCZ samples. This strong phenotypic association across ASD and SCZ offers a basis to study the known genetic and neurocognitive overlap between the two diagnoses.^44^

The A3 dimension was associated with a high performance IQ and a high ADOS score: trends opposite to the A1 dimension. While individuals with ASD, under the same umbrella, have been found to show impaired cognition at the group level, a significant heterogeneity has been noted with reports suggesting potential clusters based on IQ.^45^ Although not a formal category in the current diagnostic manuals of ASD, approximately two thirds of individuals on the autism spectrum have a normal to high IQ.^46^ Investigating individuals with high A3 scores separately may better explain this distinctive phenomenon.

### 3.3. Genetic architecture of the three endophenotypic dimensions in the general population

The three ASD dimensions were significantly heritable: 38-71% SNP-based heritability estimates. This implicated the genetic underpinnings of the neuroanatomical endophenotypes – premised on heritability – associated with ASD in the general population. Comorbidities with other psychiatric or mood disorders, such as SCZ and anxiety disorders in the UK Biobank population, might partially account for expressing the three dimensions due to overlapping genetic architecture. ASD is highly heritable, with most estimates ranging from 50–90% in the literature.^47^ Our dimensional representation may highlight potentially high-risk individuals in the general population without being clinically diagnosed with ASD.

The three ASD dimensions demonstrated distinct genetic associations at the SNP, gene, and gene-set pathway levels. The A1 dimension was identified with one novel genomic locus, with its candidate SNPs being previously associated with various imaging-derived measures (e.g., brain volume).^48^ None of the identified genes were associated with ASD, but the *FOXO3* gene is a well-known factor of human longevity and aging.^49^ The A1 dimension was positively correlated with the S1 dimension and negatively with S2: confluent evidence from both genetic (***Fig. 4d***) and imaging-dimensional correlations (***Fig. 2b***). Taken together, we speculate that this “aging-related” A1 dimension is driven by individuals with an ASD behavioral phenotype profile who also suffer non-ASD-specific brain atrophy associated with advanced brain aging.

The A2 dimension was associated with one novel genomic locus and one candidate SNP (rs1127905) that was associated with SCZ.^50^ Moreover, it was neuroanatomically and genetically correlated with both S1 and S2 dimensions. Given also its association with antipsychotic medication use, we speculate that this “schizophrenia-like” A2 dimension is driven by individuals with a high risk of developing psychiatric conditions as adults, while exhibiting autistic symptoms at younger ages. The significant correspondence between A2 and S2 with their shared imaging patterns, clinical profiles, and genetic architecture indicate a possibility of identifying common biological mechanisms across these mental health diagnoses.^51^ As of October of 2021, 719 unique SNPs have been mapped to “Autism Spectrum Disorder” on the GWAS Catalog,^32^ while 573 (80%) of them have also been mapped to “Schizophrenia”.

One of the associated genes of the A2 dimension (*DEFB119*) was previously associated with the pathway of the innate immune system in individuals with ASD.^52^ Furthermore, the A2 dimension was overrepresented in the biological pathway of hydrogen peroxide-mediated programmed cell death. Many studies have shown that oxidative stress, partly due to hydrogen peroxide (H_2_O_2_), is elevated in individuals with ASD.^53^ ASD has been considered more than a brain condition. For example, its links to the immune system or nervous system abnormalities have been highlighted.^10^ Genes associated with the A2 dimension demonstrated stable, high expression in cerebellum and other nervous tissues, supporting the hypothesis that the effects of ASD extend beyond the brain.

The lead SNP (rs72810098) of the A3 dimension was previously associated with the sex hormone-binding globulin levels.^54^ The *C20orf112* gene was speculated to be involved in brain development. This dimension implicated the biological pathway regulating the intrinsic apoptotic signaling pathway. Programmed cell/neuron death (i.e., apoptosis) is critical to influencing the morphology of the human brain by disconnecting the proper neural wiring and a possible connection between neural cell death and ASD has been demonstrated.^55^ In addition, the synaptic formation was found to be disrupted in individuals with ASD.^56^ Lastly, our genetic correlation result – A3 is positively correlated with ASD – supports the claim that the A3 dimension represents the “classical ASD” mechanism. Our genetic analyses complement the imaging and clinical findings of characterizing the heterogeneity of the three ASD dimensions, supporting the well-known “Cheverud’s Conjecture” – the observation that genetic correlations usually mirror phenotypic correlations.^57^

### 3.4. Limitations and Future Work

ASD is more prevalent in males, with a male-to-female ratio of around four.^2^ Consequently, in this study, only 12% of individuals with ASD and 15% of TD controls were females. However, the CV showed that the ASD models trained with only male participants still reliably predicted the dimension scores for the females, supporting the effectiveness of the sex-wise harmonization.

For 34% of the individuals with ASD (*N*=104), all three ASD dimension scores were less than zero, indicating that their brains were not significantly different than the TD controls. Neurodevelopmentally, their structural brains have either been normal since birth, or have converged to look similar to those of the TD controls at the time of their scan. To better characterize this considerable proportion of ASD, we may need to seek other imaging modalities or investigate their longitudinal developmental trends.

In summary, this work characterizes the brain structural heterogeneity within the late adolescent to adult ASD population by offering an endophenotypic three-dimensional representation with three novel neuroanatomical signatures. These findings pave an important research avenue for understanding the intricate variations and diverse underlying etiological mechanisms within the ASD population.

## 4. Methods

### 4.1. Study sample and image acquisition

A demographic summary of the samples used in this study is presented in ***Table 1***. Analyzed imaging data included T1-weighted brain MRI of 307 individuals with ASD (25.4±9.8 years, 273 males) and 362 age- and sex-matched TD controls (25.8±8.9 years, 309 males) between the ages of 16 and 64 shared by the ABIDE (Autism Brain Imaging Data Exchange) consortium.^24^ Only data from 16 sites with more than 10 controls within the age range were included to ensure robust site-wise image harmonization, as outlined in 4.3. A summary of the 16 sites can be found in ***Supplementary Table 7***.

For comparison, data from the PHENOM consortium that were used to derive neuroanatomical dimensions of SCZ^21^ were also investigated: 307 individuals with SCZ (30.9±7.3 years, 199 males) and 364 TD controls (29.5±7.0 years, 203 males). Additionally, 18,678 individuals (57.4±4.3 years, 8,015 males) from the UK Biobank cohort^25^ representing the general population were investigated. A broad range of disease diagnoses based on the clinical history was also provided (https://biobank.ctsu.ox.ac.uk/crystal/coding.cgi?id=19&nl=1). All participants recruited in the above studies provided informed written consent according to the protocol approved by each of their Institutional Review Boards.

### 4.2. Image preprocessing and quality assurance

All raw T1-weighted MRI (182×218×182 voxels) from the ABIDE (*N*=778 individuals initially) were manually examined for head motion, image artifacts, or restricted field-of-view. Two individuals were excluded due to significant noise in the images. Images were then corrected for magnetic field inhomogeneity, and a multi-atlas segmentation^58^ was applied to obtain brain volumes in 145 anatomical ROIs (***Supplementary Table 1***). Two individuals were further excluded due to under-segmentation. The final sample (*N*=669) after matching the two groups for mean age and sex has been outlined above. The MR images of PHENOM and UK Biobank followed the same quality assurance procedure and were detailed elsewhere.^21,36^

### 4.3. Image harmonization

Site-wise differences in the 145 ROI volumes were linearly harmonized using ComBat^59^, while preserving effects of age, sex, and ICV, based on the control group of the ABIDE sample. For the control-based site-wise harmonization purposes, only cognitively normal UK Biobank participants without the history of depression (*N*=1,510), diabetes mellitus (*N*=742), stroke (*N*=123), bipolar disorder (*N*=60), or any mental, behavioral, neurodevelopmental disorders (code F01-F99 from the ICD-10;^26^ *N*=1,554) were treated as the reference control group (*N*=15,325). Subsequently, the ROI volumes were linearly adjusted for age- and sex-wise differences, based on the trends in the TD group. For every validation or test dataset (PHENOM or UK Biobank), to prevent information leakage, its harmonization was performed separately and based on the TD group of the training dataset (on ABIDE). Then the corrected ROIs were normalized to *z*-scores and were used as features in the HYDRA models. ***Supplementary Fig. 11*** contains a flowchart to summarize the harmonization procedure. The efficacy of the harmonization was manually checked post hoc, and the final dimension scores of the TD controls showed insignificant site-wise variations (*P*>0.9, one-way ANOVA) (***Supplementary Fig. 12***)

### 4.4. Finding neuroanatomical dimensions with HYDRA

Details of the HYDRA model can be found in Varol et al.^17^ In essence, a HYDRA model attempts to find a pre-defined number of linear support vector machine (SVM) hyperplanes (i.e., the polytope) that together enclose the control distribution while maximally separating the control group from the affected group (ASD in this case). Then, each hyperplane can be considered as a plane that is orthogonal to a dimension in which the affected group deviates from the control distribution, and projections of the data points onto this dimension can be quantified (denoted here as a “dimension score”).

The HYDRA model implemented in MATLAB R2020a was trained with the following default parameters: 10-fold nested cross-validation, initialization with determinantal point processes (DPP), L1 regularization, *C*=0.25 (regularization parameter). The number of dimensions/clusters (*k*) was varied between two and eight to find the optimal dimensional space to capture the heterogeneity of ASD neuroanatomy. As the estimated solution may differ depending on the initialization, each nested model was trained 50 times for the main model, and the derived dimension scores were averaged.

To evaluate the stability of the trained models, a metric known as the adjusted Rand index (ARI)^27^ was used. ARI is a measure of agreement between partitions and is frequently used in validation of clustering analyses for its relative insensitivity to the number of clusters. ARI was computed per HYDRA model by treating the neuroanatomical dimensions as clusters (i.e., a subject is assigned to a cluster corresponding to the dimension with the largest score received). To test the significance of each ARI measure, a permutation test was followed with the group labels (ASD versus TD) shuffled. This was repeated 100 times to build the “null distribution” of the ARI, and *t*-tests were performed to compute significance. Both a high ARI and a significant difference from the null distribution were desired in a stable HYDRA model.

A set of thorough cross-validations were also performed. First, leave-one-site-out cross-validation was conducted for the three largest sites. Second, all female participants (*N*=87) were held out during the training and used in the testing. Third, a stratified 10-fold cross-validation was performed with stratification by site, sex, and diagnosis. Fourth, a stratified 2-fold (split-half) cross-validation was performed for the model with the optimal *k* to verify that the neuroanatomical patterns captured by the dimensions were reproducible. Each cross-validated model was trained 20 times with different random initialization. Pairwise mean absolute error (MAE) between dimension scores was computed.

The ASD dimension scores from the stratified 10-fold cross-validation were used as the final scores for the individuals of ABIDE. The final model that was trained with the full ABIDE dataset was applied to compute the ASD dimension scores for the PHENOM and UK Biobank participants. The raw dimension scores were adjusted for ICV using association parameters estimated in the control group. Both sets of the raw and the ICV-corrected dimension scores were investigated.

### 4.5. Characterizing anatomical patterns of ASD dimensions

The T1-weighted images were transformed to the voxel-based RAVENS (Regional Analysis of Volumes Examined in Normalized Space) maps^60^ for gray and white matter and were harmonized using the same methods as with the ROIs. Pearson correlations between the final ASD dimension scores and the harmonized RAVENS maps were computed and the *P*-values were adjusted with the Bonferroni correction. Additionally, the dimension scores were correlated to the brain age gap (predicted age based on neuroanatomy minus the actual chronological age)^30^ for the individuals of UK Biobank.

### 4.6. Associations to clinical variables and other diagnoses

ASD dimension scores were correlated with the ADOS scores^29^ and three types of IQ (full-scale, verbal, and performance). The majority of the ABIDE participants (>82%) received variations of the Wechsler IQ tests.^28^ The correlation was computed first with all IQ scores, and then with only scores from the Wechsler tests to confirm whether the same trends were observed (***Supplementary Table 3***). The dimension scores were also related to the medication status for individuals with ASD. Out of 203 individuals who reported their medication status, 61 (30.1%) were currently taking related medication (within three months from the MRI scan). ***Supplementary Table 4*** provides the complete list of medications. A complete list of demographic and clinical variables investigated, including those from PHENOM and UK Biobank datasets, are summarized in ***Supplementary Table 2***. All *P*-values were adjusted for multiple comparisons using the Benjamini-Hochberg false discovery rate correction when establishing significance.

### 4.7. Genetic quality check protocol

The genetic data for the UK Biobank participants (version 3) were downloaded from their website (https://www.ukbiobank.ac.uk/enable-your-research/about-our-data/genetic-data) on July 2021. We used the imputed genetic data in the current study. Details regarding the imputation can be found in the original UK Biobank paper.^25^ The genotyped and imputed SNPs (single nucleotide polymorphisms) from the UK Biobank were preprocessed with a standard quality check (QC) protocol (https://www.cbica.upenn.edu/bridgeport/data/pdf/BIGS_genetic_protocol.pdf). ^36^

First, we extracted and excluded related individuals (2nd-degree related individuals) in the complete UK Biobank sample (*N*=488,377) via King software relationship inference.^61^ Duplicated variants from all 22 autosomal chromosomes were then removed. Subsequently, we consolidated datasets for participants with both imaging and genetics data to have both modalities. Participants were removed if they had unmatched sex identities between genetically-identified sex and self-acknowledged sex. Lastly, subjects with i) more than 3% of missing genotypes, ii) variants with a minor allele frequency of less than 1%, iii) variants with larger than 3% of the missing genotyping rate, or iv) variants that failed the Hardy-Weinberg test at 1×10^−10^ were removed. Finally, 33,541 participants and 8,469,833 SNPs passed the QC procedures to be used for the subsequent genome analyses. To adjust for population stratification,^62^ we derived the first 40 genetic principal components (PC) using the FlashPCA software.

### 4.8. Genome-wide associations

After the QC, 14,786 participants with European ancestry and within the age range of the ABIDE sample were filtered. The three ASD dimension scores and two SCZ dimension scores were treated as phenotypes of interest and were fit into multiple linear regression models for GWAS using Plink 2 (v2.0.0).^63^ Age (at the time of scan), age-squared, sex, age-sex interaction, age-squared-sex interaction, ICV, and the first 40 genetic principal components (PC) were used as covariates. The FUMA online platform (v1.3.7)^34^ was then used to annotate the genomic risk loci, lead SNPs, candidate SNPs, and independent significant SNPs (***Supplementary Notes 1***). For the lead SNPs identified in our GWAS, we then manually queried them in the GWAS Catalog.^32^ Lead SNPs that had not been associated with any clinical traits in the GWAS Catalog were labeled as novel. GCTA-GREML^64^ was used to estimate the SNP-based heritability of the dimension scores in the general population, while the same confounders used in GWAS were adjusted for. One-sided likelihood ratio tests were performed to derive the *P*-values.

### 4.9. Gene-level associations and gene set pathway enrichment

MAGMA (v1.0.8)^33^ was used for gene-level associations. Gene annotation was performed to map the valid SNPs (reference variant location from the Phase 3 of 1,000 Genomes for European ancestry) to genes (human genome build 37). The gene-level associations used the SNP-level GWAS summary statistics to obtain gene-level *P*-values between each phenotype and the 18,902 protein-encoding genes. In addition, gene set enrichment analyses were conducted using gene sets from the MsigDB database (v6.2),^65^ including 4,761 curated gene sets and 5,917 ontology gene sets. Bonferroni correction was performed for all tested genes (*P*<2.64×10^−6^) and gene sets (*P*<4.68×10^−6^). All other parameters were set by default in FUMA.

### 4.10. Gene expressions

FUMA provides a core functionality (*GENE2FUNC*) to analyze gene expression in the phenotype of interest (therein A1, A2, and A3). In particular, *GENE2FUNC* took the prioritized, mapped genes (by default in FUMA) as input. The gene expression heat maps were generated using GTEx version 8 (54 tissue types and 30 general tissue types).^66^ The average expression per label (log_2_ transformed in each tissue type) was displayed on the corresponding heat maps. All other parameters were set by default in FUMA.

### 4.11. Genetic Correlations

LDSC^35^ was used to estimate the pairwise genetic correlation between each dimension score and seven neurodegenerative and neuropsychiatric diseases, including Alzheimer’s disease (AD), attention-deficit/hyperactivity disorder (ADHD), ASD, bipolar disorder, major depressive disorder (MDD), SCZ, OCD (obsessive-compulsive disorder). The details of the inclusion criteria were described elsewhere.^36^ LDSC uses the GWAS summary statistics from our study and previous studies for the seven selected diseases to determine the proportion of variance that two clinical traits share due to genetic causes.

## Data Availability

The ABIDE dataset is available from the National Institute of Mental Health Data Archive (NDA) upon permission (https://nda.nih.gov/edit_collection.html?id=2039). The UK Biobank dataset is available from their website upon permission (https://biobank.ndph.ox.ac.uk/ukb/). The dimension scores for both ASD and SCZ that support the findings of this study are available upon reasonable request. The GWAS summary statistics are available in the BRIDGEPORT web portal: https://www.cbica.upenn.edu/bridgeport/. The MsigDB database for gene-set enrichment analysis is available at: https://www.gsea-msigdb.org/gsea/msigdb/.

## 6. Code Availability

The software and resources developed by our team or by other research groups used in this study are all publicly available:

- HYDRA: Matlab implementation: https://github.com/evarol/HYDRA, Python implementation: https://github.com/anbai106/mlni
- Genetic quality check protocol: https://www.cbica.upenn.edu/bridgeport/data/pdf/BIGS_genetic_protocol.pdf
- FUMA: https://fuma.ctglab.nl/
- MUSE for image segmentation: https://www.med.upenn.edu/sbia/muse.html
- PLINK for GWAS: https://www.cog-genomics.org/plink/
- GCTA for heritability estimates: https://yanglab.westlake.edu.cn/software/gcta/
- LDSC for genetic correlation estimates: https://github.com/bulik/ldsc
- MAGMA for gene analysis: https://ctg.cncr.nl/software/magma

## 7. Acknowledgements

The authors would like to thank the investigators of ABIDE, PHENOM, and UK Biobank, as well as all the participants and their families. This work was supported in part by NIH grant R01MH112070 and R01MH123550.

## 8. Author Contributions

Dr. Hwang, Dr. Wen, and Dr. Sotardi take full responsibility for the integrity of the data and the accuracy of the data analysis.

*Study concept and design*: Hwang, Wen, Sotardi, Davatzikos

*Acquisition, analysis, or interpretation of data*: Hwang, Wen, Sotardi, Davatzikos

*Drafting of the manuscript*: Hwang, Wen, Sotardi, Davatzikos

*Critical revision of the manuscript for important intellectual content*: all authors

*Statistical analysis*: Hwang

*Genetic analysis*: Wen

*Study supervision*: Davatzikos

## 9. Ethics Declarations

No competing interests to report.

## Notes

### Competing Interest Statement

The authors have declared no competing interest.

### Author Declarations

The study used ONLY openly available human data from three studies or consortia: ABIDE, PHENOM, and UK Biobank. The ABIDE dataset is available from the National Institute of Mental Health Data Archive (NDA) upon permission (https://nda.nih.gov/edit_collection.html?id=2039). The UK Biobank dataset is available from their website upon permission (https://biobank.ndph.ox.ac.uk/ukb/). The dimension scores for both ASD and SCZ that support the findings of this study are available from the authors of the originating paper upon reasonable request (https://pubmed.ncbi.nlm.nih.gov/32103250/). All participants recruited in the above studies provided informed written consent according to the protocol approved by each of their Institutional Review Boards.

