## Supplementary Material for "Three Imaging Endophenotypes Characterize Neuroanatomical Heterogeneity of Autism Spectrum Disorder"

### Supplementary Information for "Three Imaging Endophenotypes Characterize Neuroanatomical Heterogeneity of Autism Spectrum Disorder"

Gyujoon Hwang[1,*], Junhao Wen[1,*], Susan Sotardi[2,*], Edward S. Brodkin[3], Ganesh B. Chand[1,4], Dominic B. Dwyer[5], Guray Erus[1], Jimit Doshi[1], Pankhuri Singhal[6], Dhivya Srinivasan[1], Erdem Varol[1,7], Aristeidis Sotiras[1,4], Paola Dazzan[8], Rene S. Kahn[9], Hugo G. Schnack[10], Marcus V. Zanetti[11,12], Eva Meisenzahl[13], Geraldo F. Busatto[11], Benedicto Crespo-Facorro[14], Christos Pantelis[15], Stephen J. Wood[16], Chuanjun Zhuo[17], Russell T. Shinohara[1,18], Haochang Shou[1,18], Yong Fan[1], Nikolaos Koutsouleris[5], Raquel E. Gur[3], Ruben C. Gur[3], Theodore D. Satterthwaite[1],[3], Daniel H. Wolf[1,3], Christos Davatzikos[1]

[*]*These authors contributed equally to this work*

[1]*Center for Biomedical Image Computing and Analytics, Perelman School of Medicine, University of Pennsylvania, Philadelphia, USA*
[2]*Department of Radiology, Children's Hospital of Philadelphia, Philadelphia, USA*
[3]*Department of Psychiatry, Perelman School of Medicine, University of Pennsylvania, Philadelphia, USA*
[4]*Department of Radiology, School of Medicine, Washington University in St. Louis, St. Louis, USA*
[5]*Department of Psychiatry and Psychotherapy, Ludwig-Maximilian University, Munich, Germany*
[6]*Department of Genetics, Perelman School of Medicine, University of Pennsylvania, Philadelphia, USA*
[7]*Department of Statistics, Zuckerman Institute, Columbia University, New York, USA*
[8]*Department of Psychological Medicine, Institute of Psychiatry, Psychology and Neuroscience, King's College London, London, UK*
[9]*Department of Psychiatry, Icahn School of Medicine at Mount Sinai, New York, USA*
[10]*Department of Psychiatry, University Medical Center Utrecht, Utrecht, Netherlands*
[11]*Institute of Psychiatry, Faculty of Medicine, University of São Paulo, São Paulo, Brazil*
[12]*Hospital Sírio-Libanês, São Paulo, Brazil*
[13]*LVR-Klinikum Düsseldorf, Kliniken der Heinrich-Heine-Universität, Düsseldorf, Germany*
[14]*University Hospital Virgen del Rocio, Department of Psychiatry, School of Medicine, IBiS-CIBERSAM, University of Sevilla, Spain*
[15]*Melbourne Neuropsychiatry Centre, Department of Psychiatry, University of Melbourne and Melbourne Health, Carlton South, Australia*
[16]*Orygen, Melbourne, Australia; Centre for Youth Mental Health, University of Melbourne, Melbourne, Australia; School of Psychology, University of Birmingham, Edgbaston, UK*
[17]*Department of Psychiatric-Neuroimaging-Genetics and Co-morbidity Laboratory, Tianjin Anding Hospital; Department of Psychiatry, Tianjin Medical University, Tianjin, China*
[18]*Penn Statistics in Imaging and Visualization Center, Department of Biostatistics, Epidemiology, and Informatics, Perelman School of Medicine, University of Pennsylvania, Philadelphia, USA*

**Table S1.** 145 whole-brain anatomical regions of interest (ROIs)[1] investigated

| | | |
|---|---|---|
| 3rd ventricle | Anterior insula (R) | Occipital pole (R) |
| 4th ventricle | Anterior insula (L) | Occipital pole (L) |
| Accumbens area (R) | Anterior orbital gyrus (R) | Occipital fusiform gyrus (R) |
| Accumbens area (L) | Anterior orbital gyrus (L) | Occipital fusiform gyrus (L) |
| Amygdala (R) | Angular gyrus (R) | Opercular part of inferior frontal gyrus (R) |
| Amygdala (L) | Angular gyrus (L) | Opercular part of inferior frontal gyrus (L) |
| Brain Stem | Calcarine cortex (R) | Orbital part of inferior frontal gyrus (R) |
| Caudate (R) | Calcarine cortex (L) | Orbital part of inferior frontal gyrus (L) |
| Caudate (L) | Central operculum (R) | Posterior cingulate gyrus (R) |
| Cerebellum exterior (R) | Central operculum (L) | Posterior cingulate gyrus (L) |
| Cerebellum exterior (L) | Cuneus (R) | Precuneus (R) |
| Cerebellum WM (R) | Cuneus (L) | Precuneus (L) |
| Cerebellum WM (L) | Entorhinal area (R) | Parahippocampal gyrus (R) |
| Hippocampus (R) | Entorhinal area (L) | Parahippocampal gyrus (L) |
| Hippocampus (L) | Frontal operculum (R) | Posterior insula (R) |
| Inferior lateral ventricle (R) | Frontal operculum (L) | Posterior insula (L) |
| Inferior lateral ventricle (L) | Frontal pole (R) | Parietal operculum (R) |
| Lateral ventricle (R) | Frontal pole (L) | Parietal operculum (L) |
| Lateral ventricle (L) | Fusiform gyrus (R) | Postcentral gyrus (R) |
| Pallidum (R) | Fusiform gyrus (L) | Postcentral gyrus (L) |
| Pallidum (L) | Gyrus rectus (R) | Posterior orbital gyrus (R) |
| Putamen (R) | Gyrus rectus (L) | Posterior orbital gyrus (L) |
| Putamen (L) | Inferior occipital gyrus (R) | Planum polare (R) |
| Thalamus proper (R) | Inferior occipital gyrus (L) | Planum polare (L) |
| Thalamus proper (L) | Inferior temporal gyrus (R) | Precentral gyrus (R) |
| Ventral diencephalon (R) | Inferior temporal gyrus (L) | Precentral gyrus (L) |
| Ventral diencephalon (L) | Lingual gyrus (R) | Planum temporale (R) |
| Cerebellar vermal lobules I-V | Lingual gyrus (L) | Planum temporale (L) |
| Cerebellar vermal lobules VI-VII | Lateral orbital gyrus (R) | Subcallosal area (R) |
| Cerebellar vermal lobules VIII-X | Lateral orbital gyrus (L) | Subcallosal area (L) |
| Basal forebrain (R) | Middle cingulate gyrus (R) | Superior frontal gyrus (R) |
| Basal forebrain (L) | Middle cingulate gyrus (L) | Superior frontal gyrus (L) |
| Frontal lobe WM (R) | Medial frontal cortex (R) | Supplementary motor cortex (R) |
| Frontal lobe WM (L) | Medial frontal cortex (L) | Supplementary motor cortex (L) |
| Occipital lobe WM (R) | Middle frontal gyrus (R) | Supramarginal gyrus (R) |
| Occipital lobe WM (R) | Middle frontal gyrus (L) | Supramarginal gyrus (L) |
| Parietal lobe WM (R) | Middle occipital gyrus (R) | Superior occipital gyrus (R) |
| Parietal lobe WM (L) | Middle occipital gyrus (L) | Superior occipital gyrus (L) |
| Temporal lobe WM (R) | Medial orbital gyrus (R) | Superior parietal lobule (R) |
| Temporal lobe WM (L) | Medial orbital gyrus (L) | Superior parietal lobule (L) |
| Fornix (R) | Postcentral gyrus medial segment (R) | Superior temporal gyrus (R) |
| Fornix (L) | Postcentral gyrus medial segment (L) | Superior temporal gyrus (L) |
| Anterior limb of internal capsule (R) | Precentral gyrus medial segment (R) | Temporal pole (R) |
| Anterior limb of internal capsule (L) | Precentral gyrus medial segment (L) | Temporal pole (L) |
| Posterior limb of internal capsule including cerebral peduncle (R) | Superior frontal gyrus medial segment (R) | Triangular part of the inferior frontal gyrus (R) |
| Posterior limb of internal capsule including cerebral peduncle (L) | Superior frontal gyrus medial segment (L) | Triangular part of the inferior frontal gyrus (L) |
| Corpus callosum | Middle temporal gyrus (R) | Transverse temporal gyrus (R) |
| Anterior cingulate gyrus (R) | Middle temporal gyrus (L) | Transverse temporal gyrus (L) |
| Anterior cingulate gyrus (L) | | |

* L: left hemisphere; R: right hemisphere; WM: white matter

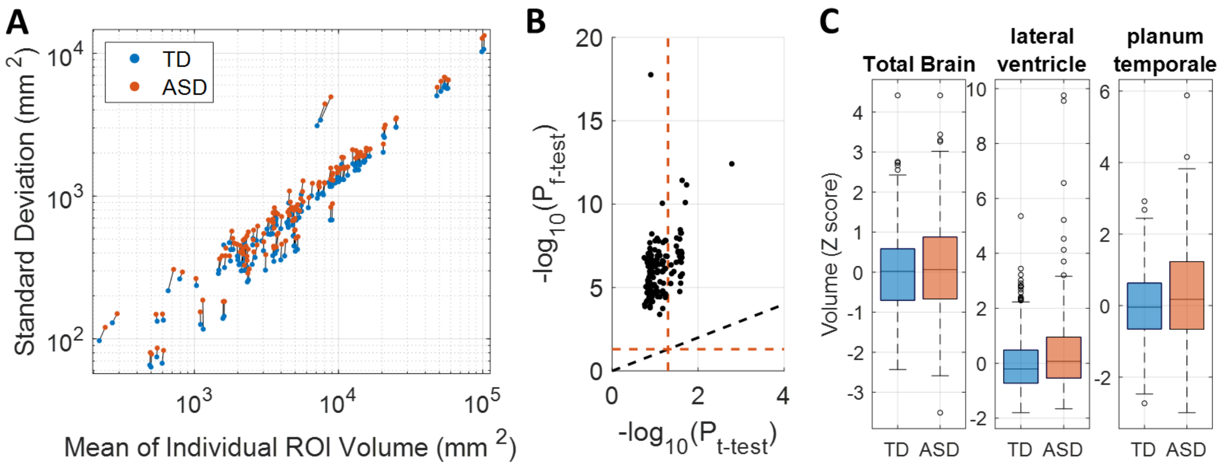


**Fig. S1: Heterogeneous Neuroanatomy of Autism Spectrum Disorder (ASD).** (A) The standard deviation and mean of brain volumes from 145 regions-of-interest (ROI) are plotted for the typically-developing (TD) (*N*=362) and ASD (*N*=307) groups. (B) FDR-corrected *P*-values from the two-sample *t*-test (without assuming equal variance) and *F*-test for equal variances between the two groups are plotted. The black dotted line locates the y=x line, and the red dotted lines mark where $P_{FDR}$=0.05. (C) Control-based *z*-scores of brain volumes are plotted for the total brain ($P_{FDR}$=0.11) and two brain regions showing the most significant ANOVA differences ($P_{FDR}$<0.04).

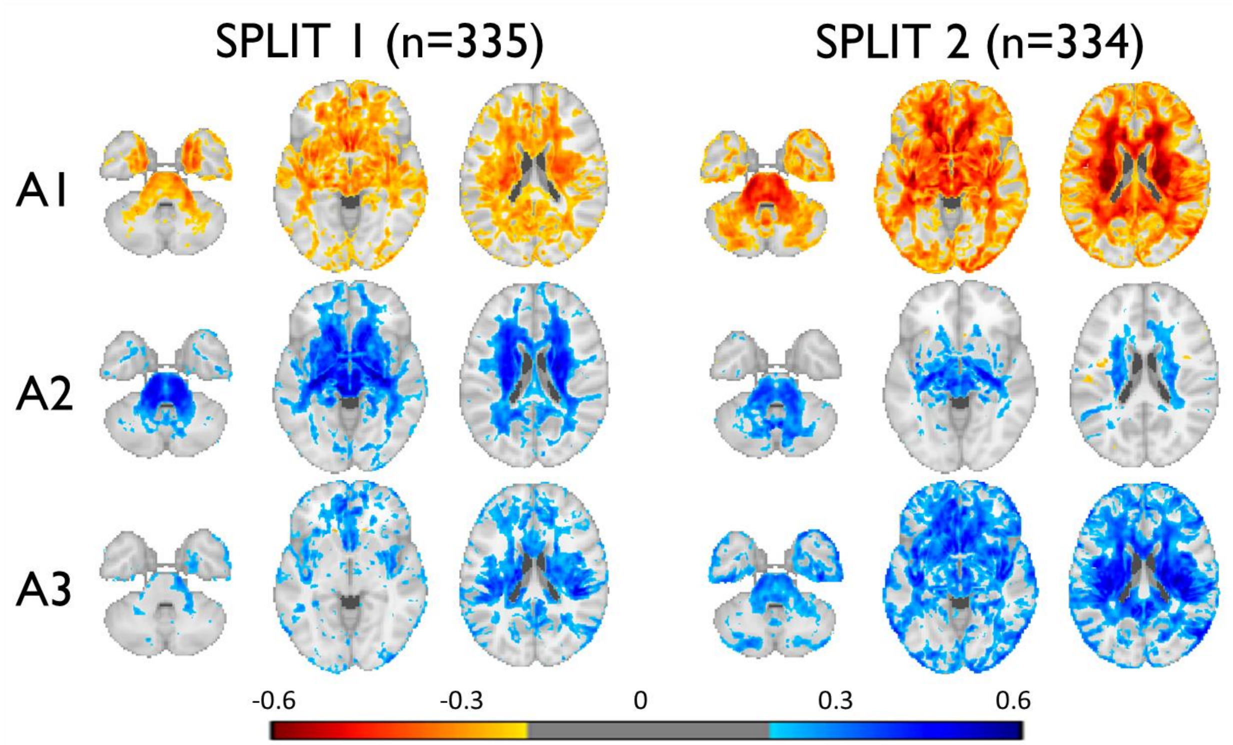


**Fig. S2: Results from the stratified 2-fold (split-half) cross-validation.** Color maps are based on the Pearson correlations between the three ASD neuroanatomical dimension scores and the voxel-wise RAVENS statistics[2] using the other left-out half, masked by abs($r$)>0.2. High pairwise correspondence was observed between the dimensions despite the significant compromise in the sample size to train these HYDRA models.

**Video S1: Three-dimensional representation of the neuroanatomical heterogeneity in ASD.** 307 individuals with ASD are projected onto the final three dimensional space to capture their heterogeneous neuroanatomy. Colored dots represent individuals who displayed high expression in one of the three dimensions. Black dots that are inside and nearby the gray rectangular cuboid (all dimension scores less than zero) represent individuals who displayed neuroanatomy similar to the TD controls.

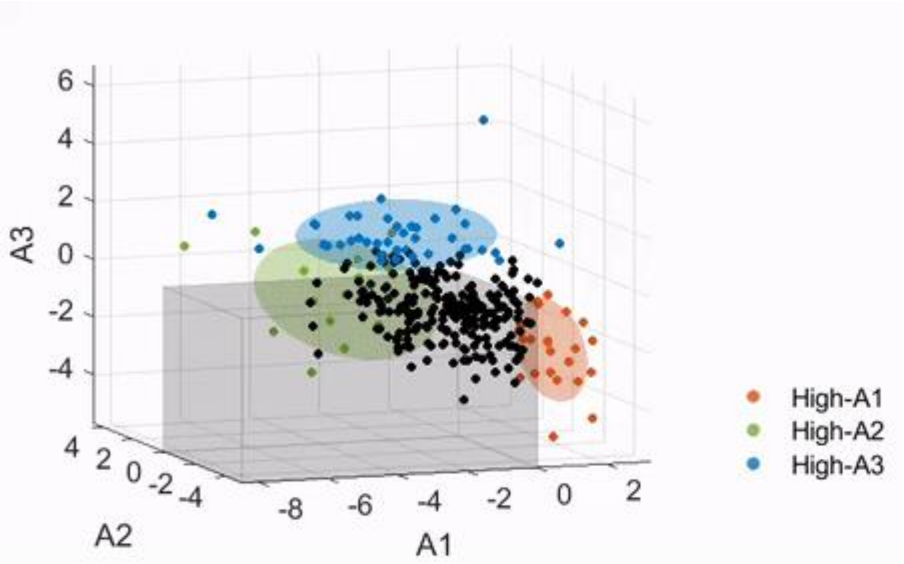

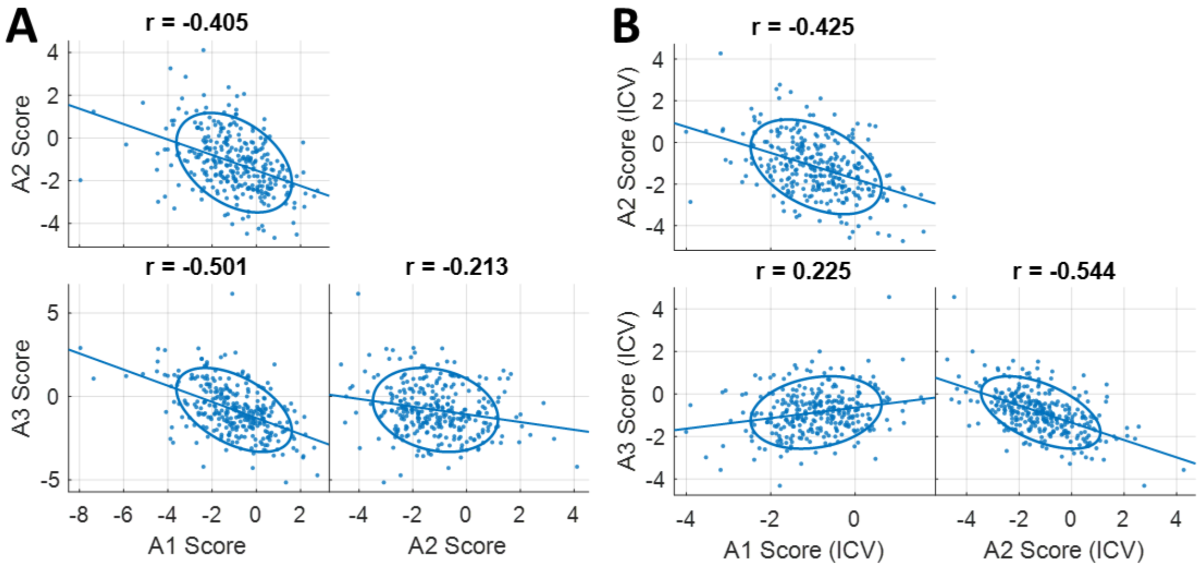


**Fig. S3: Correlations between the three ASD dimensions.** The scatter plots show relationships between the three ASD neuroanatomical dimensions among individuals with ASD (*N*=307): (A) with the raw scores, and (B) with the scores after they were adjusted for the intracranial volume (ICV).

**Table S2.** List of demographic and clinical variables investigated.

| Variable | Statistical Test | N (number of positive) | A1 | A2 | A3 | A1 (ICV) | A2 (ICV) | A3 (ICV) |
|---|---|---|---|---|---|---|---|---|
| **ABIDE (ASD)** | | | | | | | | |
| Full-scale IQ | Pearson Correlation (*r*) | 289 | -0.028 | -0.014 | 0.089 | 0.096 | -0.027 | 0.025 |
| Verbal IQ | | 231 | -0.057 | -0.008 | 0.034 | 0.033 | -0.021 | -0.044 |
| Performance IQ | | 239 | -0.135 | 0.038 | 0.124 | 0.025 | 0.002 | -0.012 |
| Performance IQ (Wechsler) | | 216 | -0.129 | 0.042 | 0.142 | 0.052 | 0.006 | 0.004 |
| ADOS Total | | 230 | -0.079 | -0.040 | 0.142 | -0.030 | -0.058 | 0.130 |
| ADOS Communication | | 217 | -0.080 | -0.040 | 0.137 | -0.054 | -0.056 | 0.137 |
| ADOS Social | | 217 | -0.101 | -0.027 | 0.131 | -0.048 | -0.045 | 0.100 |
| ADOS Stereotypical Beh. | | 211 | 0.011 | -0.022 | -0.023 | -0.043 | -0.008 | 0.018 |
| ASD Meds | Two-sample *t*-test (Cohen's *d*) | 200(12) | -0.191 | 0.321 | 0.314 | 0.115 | 0.264 | 0.166 |
| ADHD Meds | | 157(32) | -0.061 | 0.144 | 0.247 | 0.113 | 0.106 | 0.214 |
| Epilepsy/Seizure Meds | | 201(12) | 0.296 | -0.054 | -0.186 | -0.081 | 0.036 | 0.180 |
| Depression Meds | | 195(40) | -0.066 | 0.108 | -0.081 | -0.054 | 0.106 | -0.144 |
| Antipsychotics | | 203(15) | -0.201 | **0.651*** | 0.008 | -0.184 | 0.638 | -0.093 |
| OCD Meds | | 202(18) | -0.336 | 0.535 | 0.037 | -0.151 | 0.477 | -0.238 |
| Bipolar Meds | | 201(21) | 0.002 | 0.326 | 0.044 | -0.075 | 0.347 | 0.143 |
| **PHENOM (SCZ)**[3] | | | | | | | | |
| Disease Duration | Pearson Correlation (*r*) | 279 | 0.047 | -0.027 | -0.045 | -0.038 | -0.020 | 0.042 |
| Age Disease Onset | | 279 | -0.001 | 0.043 | 0.050 | 0.071 | 0.025 | 0.012 |
| PANSS-positive | | 285 | 0.017 | 0.110 | -0.028 | 0.068 | 0.110 | -0.075 |
| PANSS-negative | | 285 | 0.095 | 0.019 | -0.106 | 0.105 | 0.020 | -0.118 |
| **UK Biobank (Cognitively Normal/General Population)**[4] | | | | | | | | |
| Trail Making Test-A | Pearson Correlation (*r*) | 11956 | 0.076 | 0.005 | -0.080 | 0.012 | 0.023 | -0.025 |
| Trail Making Test-B | | 11847 | 0.090 | -0.000 | **-0.101*** | -0.003 | 0.024 | -0.021 |
| Digit Symbol Substitution | | 11975 | -0.084 | 0.039 | 0.053 | -0.029 | 0.023 | -0.007 |
| Digit Span Forward | | 12294 | **-0.104*** | 0.029 | **0.122*** | 0.004 | 0.001 | 0.028 |
| Fluid Intelligence | | 17684 | **-0.127*** | 0.040 | **0.145*** | 0.022 | 0.003 | 0.010 |
| College Education | Two-sample *t*-test (Cohen's *d*) | 18537(9485) | -0.160 | 0.104 | 0.140 | 0.032 | 0.058 | -0.049 |
| Caucasian | | 17901(14725) | -0.100 | -0.057 | 0.160 | 0.001 | -0.085 | 0.089 |
| Maternal Smoking | | 4624(1496) | 0.125 | -0.137 | -0.059 | 0.068 | -0.118 | 0.026 |
| Diabetes | | 14305(742) | **0.335*** | **-0.203*** | -0.049 | **0.370*** | -0.185 | 0.046 |
| Diabetes Meds | | 15817(127) | **0.411*** | **-0.339*** | -0.023 | **0.644*** | **-0.353*** | -0.064 |
| Hyperlipidemia | | 9168(1334) | 0.113 | -0.049 | -0.086 | 0.059 | -0.031 | -0.022 |
| Hyperlipidemia Meds | | 11593(2125) | 0.155 | -0.079 | -0.065 | 0.146 | -0.066 | -0.017 |
| Hypertension | | 9960(2745) | 0.111 | 0.036 | -0.081 | 0.121 | 0.044 | -0.075 |
| Hypertension Meds | | 13674(2710) | 0.104 | 0.043 | -0.073 | 0.152 | 0.044 | -0.105 |
| Depression | | 10882(1510) | 0.020 | -0.018 | 0.009 | 0.084 | -0.028 | -0.036 |

For Pearson correlations, Pearson *r*, and for two-sample *t*-tests, Cohen's *d* are shown. A positive *d* indicates a higher score for the positive response. The three right-most columns are with ICV-corrected dimension scores.
*$P_{FDR}$<0.05, while abs(*r*)>0.1 or abs(*d*)>0.2.
ADHD: Attention-Deficit/Hyperactivity Disorder, IQ: Intelligence Quotient, OCD: Obsessive Compulsive Disorder, PANSS: Positive and Negative Syndrome Scale

**Table S3.** List of test types used to measure intelligence quotient (IQ).

| **Test Type** | **Number of participants (ASD/TD)** | | | **Variations** |
|---|---|---|---|---|
| | **Full-scale (FIQ)** | **Verbal (VIQ)** | **Performance (PIQ)** | |
| WASI (Wechsler Abbreviated Scale Intelligence)[5] | 170/166 | 169/166 | 171/166 | WASI II |
| WAIS (Wechsler Adult Intelligence Scale)[6] | 48/60 | 25/34 | 25/34 | WAIS III, WAIS IV |
| WISC (Wechsler Intelligence Scale for Children)[7] | 19/13 | 20/17 | 20/17 | WISC III Dutch, WISC IV |
| KBIT2 (Kaufman Brief Intelligence Test 2nd Ed.)[8] | 28/29 | - | - | |
| WST (Wortschatztest)[9] | 14/27 | - | - | |
| RPM (Raven's Progressive Matrices)[10] | - | - | 13/60 | |
| PPVT (Peabody Picture Vocabulary Test)[11] | - | 7/24 | - | |
| *Average of PPVT and RPM* | *7/21* | - | - | |
| GIT (Groninger Intelligentie Test)[12] | 1/0 | 8/0 | 8/0 | |
| DAS_II (Differential Ability Scales)[13] | 2/0 | 2/0 | 2/0 | |
| **TOTAL** | **289/316** | **231/241** | **239/277** | |

**Table S4.** List of reported medications by the individuals with ASD and their classification.

| Medication | Category | Count | Variations |
|---|---|---|---|
| Methylphenidate | ADHD | 19 | Concerta(1), Metadate(1) |
| Amphetamine/ Dextroamphetamine | ADHD | 13 | Lisdexamfetamine(3) |
| Atomoxetine | ADHD | 6 | |
| Guanfacine | ADHD, Hypertension | 2 | |
| Clonidine | Hypertension | 2 | |
| Lisinopril | Hypertension | 1 | |
| Citalopram | Depression | 9 | Escitalopram(3) |
| Bupropion | Depression | 8 | |
| Venlafaxine | Depression | 4 | |
| Duloxetine | Depression | 2 | |
| Mirtazapine | Depression | 2 | |
| Trazodone | Depression | 1 | |
| **Antidepressant* | Depression | 1 | |
| Fluoxetine | Depression, OCD | 7 | |
| Sertraline | Depression, OCD | 7 | |
| Paroxetine | Depression, OCD | 2 | |
| Clomipramine | OCD | 1 | |
| Fluvoxamine | OCD | 1 | |
| Lorazepam | Epilepsy | 2 | |
| Clonazepam | Epilepsy | 1 | |
| Oxcarbazepine | Epilepsy | 1 | |
| Topiramato | Epilepsy | 1 | |
| Valproic Acid | Epilepsy, Bipolar | 6 | Divalproex(2) |
| Lamotrigine | Epilepsy, Bipolar | 2 | |
| Lithium | Bipolar | 2 | |
| Risperidone | Antipsychotics, Bipolar, ASD | 8 | |
| Aripiprazole | Antipsychotics, Bipolar, Depression, ASD | 4 | |
| Quetiapine | Antipsychotics, Bipolar, Depression | 1 | |
| Ziprasidone | Antipsychotics, Bipolar | 1 | |
| Benperidol | Antipsychotics | 1 | |

*Name of the medication not specified
ADHD: Attention-Deficit/Hyperactivity Disorder, OCD: Obsessive Compulsive Disorder

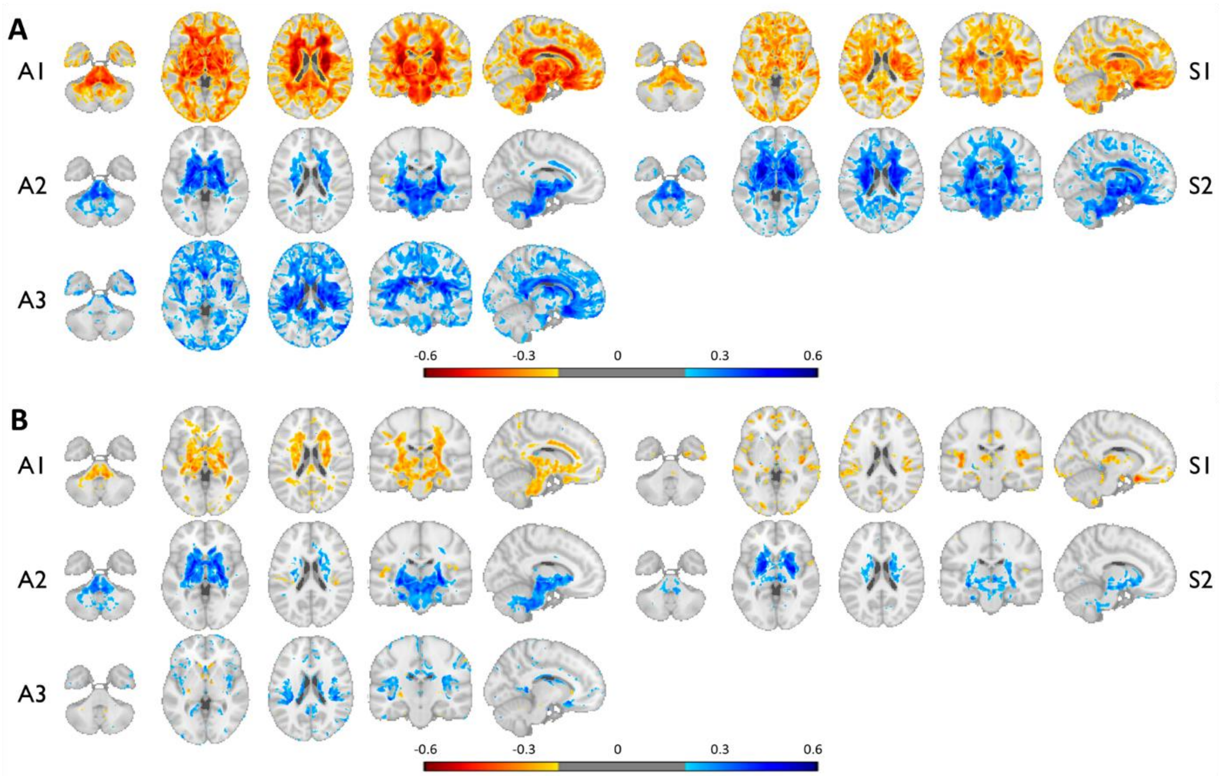


**Fig. S4: Voxel-wise patterns of ASD and SCZ dimensions.** Color maps are based on the Pearson correlations between the neuroanatomical dimension scores of ASD or SCZ and the voxel-wise RAVENS statistics using individuals with ASD ($N$=307) masked by abs($r$)>0.2. (A) Correlations are with the raw dimension scores. (B) Correlations are with the ICV-corrected dimension scores. Blue indicates increased volume with the increased dimension scores, and vice versa for the red. The A2 and S2 patterns exhibited high correspondence, while both being associated with significantly larger subcortical structures, especially the pallidum.

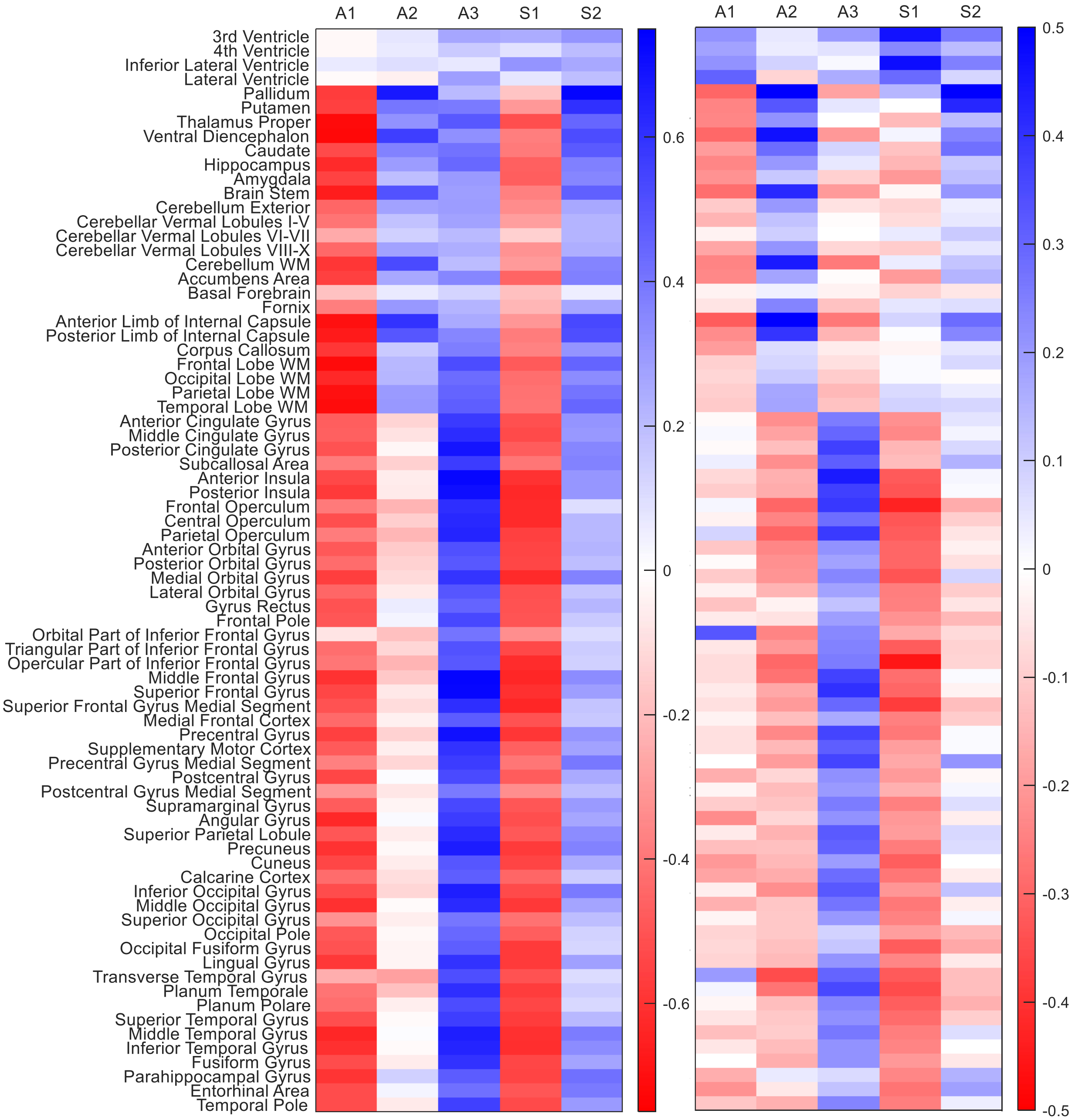


**Fig. S5: Neuroanatomical patterns captured by the dimension scores.** Color maps are based on the correlations between the dimension scores of ASD or SCZ[3] and the MUSE ROIs (left and right hemispheres merged) computed for the combined dataset of ABIDE and PHENOM ($N$=1,340). Five columns on the left are with the raw scores, and on the right are with the ICV-corrected scores.

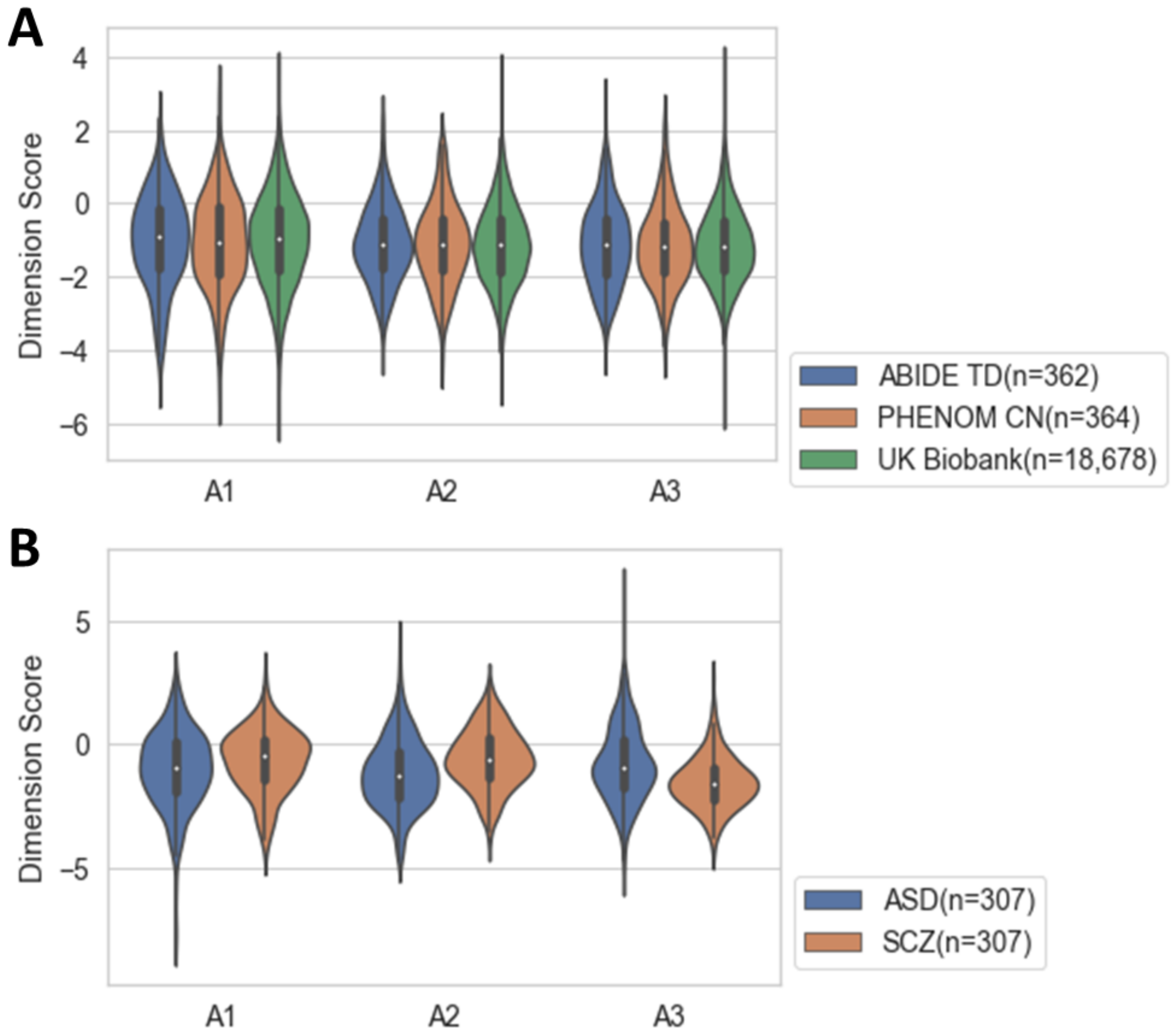


**Fig. S6: Distribution of the three dimension scores by study and diagnosis.** (A) Score distributions are plotted for the control groups per study. (B) Score distributions are plotted for individuals with ASD and SCZ.

**Table S5.** List of brain metrics investigated for the UK Biobank participants.

| Variable | N | A1 | A2 | A3 | A1 (ICV) | A2 (ICV) | A3 (ICV) |
|---|---|---|---|---|---|---|---|
| Total White Matter Lesions | 17,539 | -0.127 | 0.108 | 0.100 | -0.012 | 0.080 | -0.025 |
| SPARE-AD | 10,894 | 0.088 | 0.095 | -0.064 | 0.089 | 0.105 | -0.055 |
| SPARE-BA | 10,894 | 0.243* | 0.010 | -0.248* | 0.220* | 0.035 | -0.218* |
| Disentangled SPARE-AD | 10,894 | 0.070 | 0.142 | -0.040 | 0.070 | 0.152 | -0.027 |
| Disentangled SPARE-BA | 10,894 | 0.306* | -0.200 | -0.139 | 0.312* | -0.179 | -0.058 |

*abs($r$)>0.2, $P_{FDR}<1\times10^{-100}$

**Table S6.** Previously reported clinical associations of the two known loci
[attached separately due to large size: Supplementary_Table_6_GWAS_Catalog.csv]

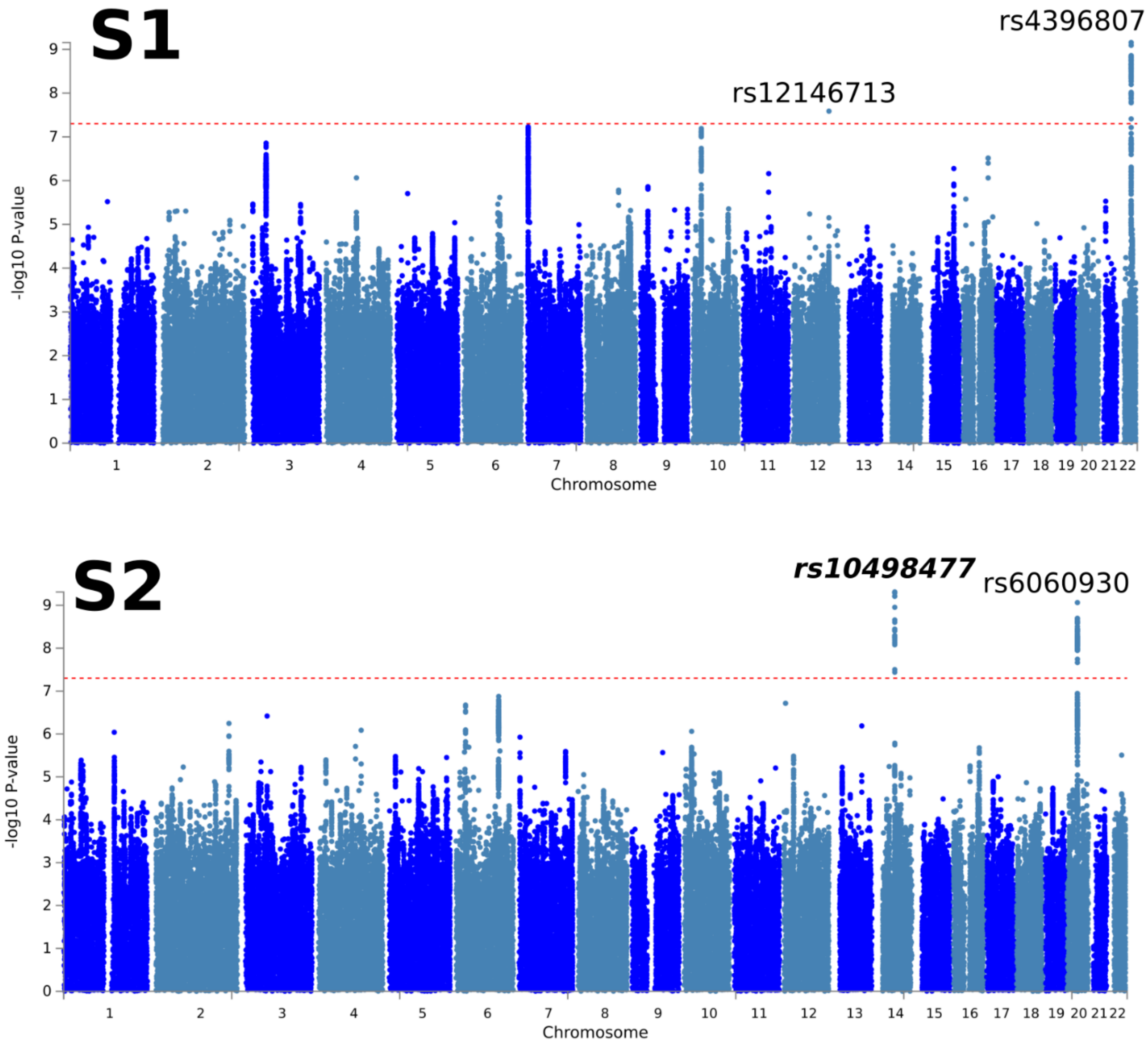


**Fig. S7**: **Heterogeneous genetic architecture of schizophrenia dimensions in the general population**. We refer to it as *de novo* (bold and italic) for the lead SNPs of all genomic risk loci if they have not been associated with any clinical traits in the GWAS Catalog. The genome-wide default *P*-value threshold ($P<5\times10^{-8}$) was used for all GWAS. S1 dimension was significantly associated with two lead SNPs (rs12146713, $P=2.58\times10^{-9}$, minor allele: C, $\beta=0.10$; rs4396807, $P=6.98\times10^{-10}$, minor allele: C, $\beta=0.07$). SNPs in these two genomic loci were previously associated with cognitive traits, brain morphology measures, etc. S2 dimension was significantly associated with two lead SNPs (rs10498477, *de novo*, $P=4.88\times10^{-10}$, minor allele: A, $\beta=-0.09$; rs6060930, $P=8.62\times10^{-10}$, minor allele: G, $\beta=-0.07$). SNPs in these two loci were previously associated with brain morphology measures.

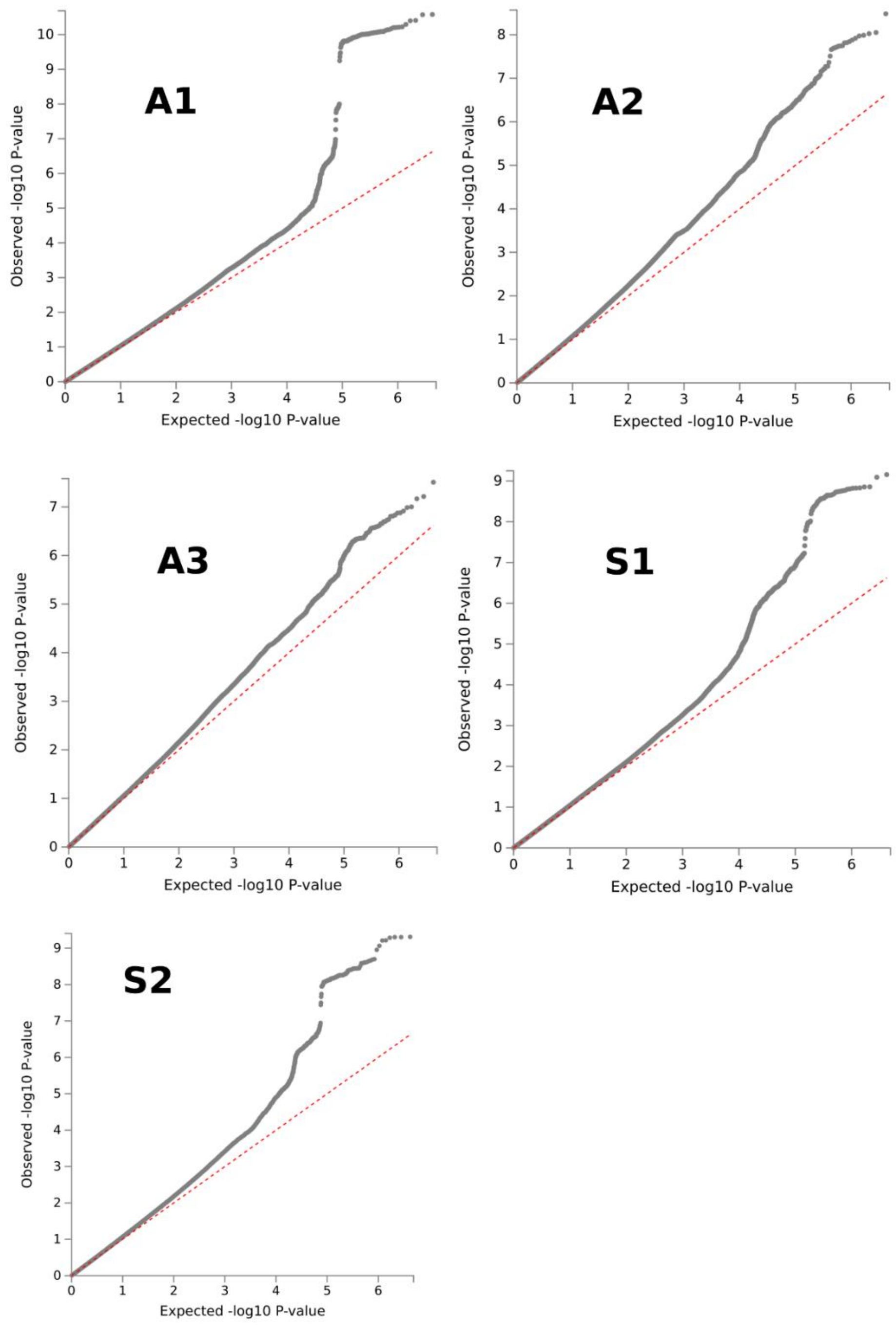


**Fig. S8**: **Quantile-quantile plots for GWAS**. QQ plots for all 5 GWAS results are presented (3 neuroanatomical dimensions of ASD and 2 dimensions of SCZ).

**Notes S1.** Definition of independent significant SNPs, lead SNPs, and genomic risk loci.

FUMA defined the significant independent SNPs, lead SNPs, candidate SNPs, and genomic risk loci as follows (https://fuma.ctglab.nl/tutorial#snp2gene):

*Independent significant SNPs*
They are defined as SNPs with $P \leq 5 \times 10^{-8}$ that are independent of each other at the user-defined $r^2$ (set to 0.6 in the current study). We further describe *candidate SNPs* as those in linkage disequilibrium (LD) with the independent significant SNPs. FUMA then queries each candidate SNP in the GWAS Catalog to check whether any clinical traits have been reported to be associated with previous GWAS studies.

*Lead SNPs*
Lead SNPs are defined as independent significant SNPs that are also independent from each other at $r^2 < 0.1$. If multiple independent significant SNPs are correlated at $r^2 \geq 0.1$, then the one with the lowest individual *P*-value becomes the lead SNP. If $r^2$ threshold is set to 0.1 for the independent significant SNPs also, then they would constitute the identical set as the lead SNPs by definition. FUMA thus advises setting $r^2$ to be 0.6 or higher.

*Genomic risk loci*
FUMA defines genomic risk loci to include all independent signals that are physically close or overlapping in a single locus. First, independent significant SNPs dependent on each other at $r^2 \geq 0.1$ are assigned to the same genomic risk locus. Then, independent significant SNPs with less than the user-defined distance (250kilobase by default) away from one another are merged into the same genomic risk locus—the distance between two LD blocks of two independent significant SNPs is the distance between the closest points from each LD block. Each locus is represented by the SNP within the locus with the lowest *P*-value.

**Table S7.** Differences in gene-level associations

| ASD Dimension | Gene | $P$ | $Z$ |
|---|---|---|---|
| A1 | *CDC20* | $1.32\times10^{-7}$ | 5.14 |
| | *MPL* | $5.21\times10^{-7}$ | 4.88 |
| | *GMNC* | $7.82\times10^{-7}$ | 4.80 |
| | *HYI* | $8.49\times10^{-7}$ | 4.78 |
| | *FOXO3* | $1.62\times10^{-6}$ | 4.65 |
| A2 | *DEFB124* | $8.60\times10^{-8}$ | 5.22 |
| | *FOXO3* | $1.59\times10^{-7}$ | 5.11 |
| | *REM1* | $1.77\times10^{-7}$ | 5.09 |
| | *PAPPA* | $3.69\times10^{-7}$ | 4.95 |
| | *DEFB119* | $5.99\times10^{-7}$ | 4.86 |
| | *STX6* | $1.24\times10^{-6}$ | 4.71 |
| | *SELO* | $1.60\times10^{-6}$ | 4.66 |
| A3 | *C20orf112* | $3.09\times10^{-7}$ | 4.98 |
| | *METTL16* | $1.33\times10^{-6}$ | 4.69 |

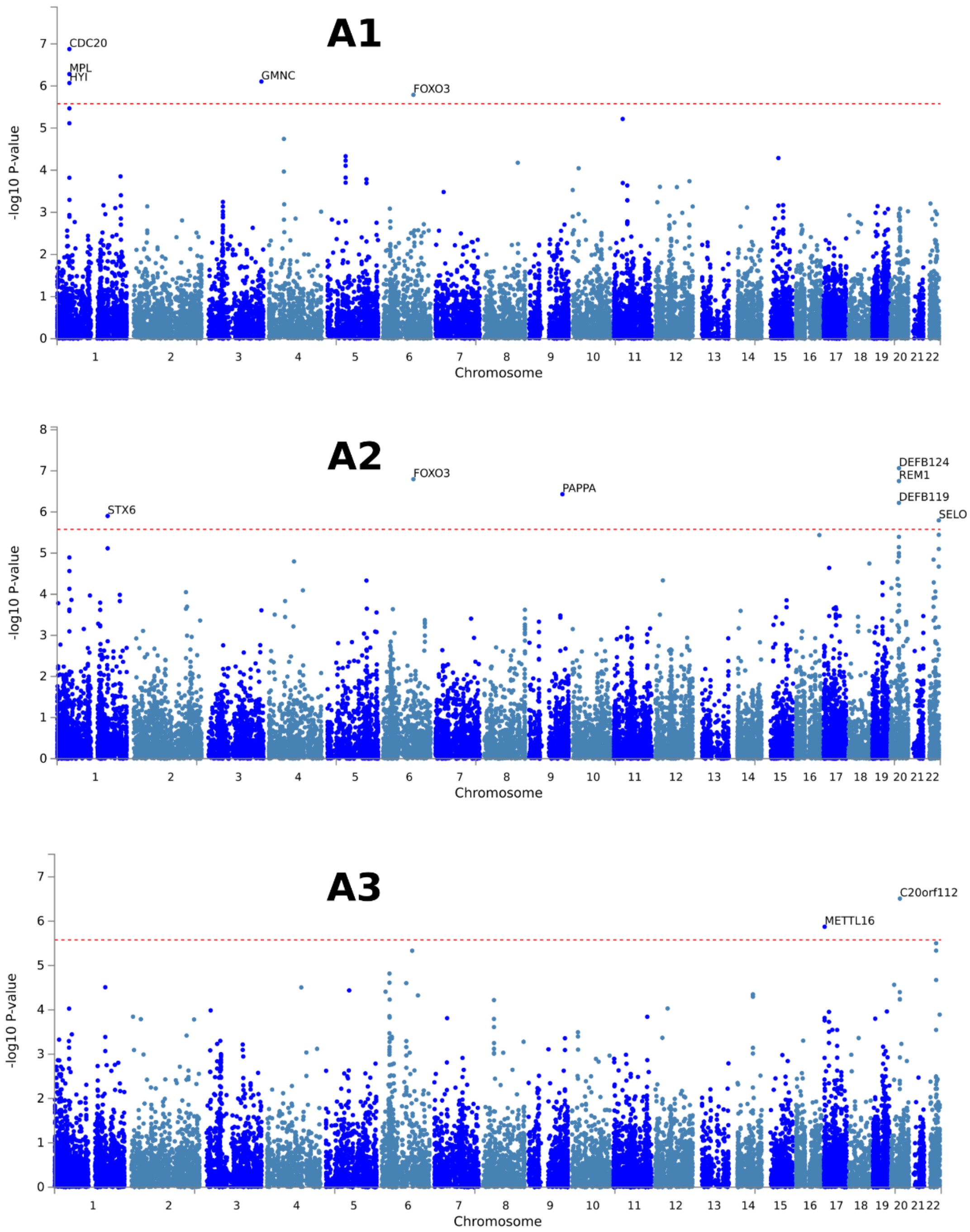


**Fig. S9**: **Gene-level associations for the ASD dimensions**. Gene-level associations were performed by MAGMA based on the input GWAS summary statistics. Input SNPs were first mapped to 18,902 protein-coding genes, and then MAGMA derived the gene-level *P*-values.

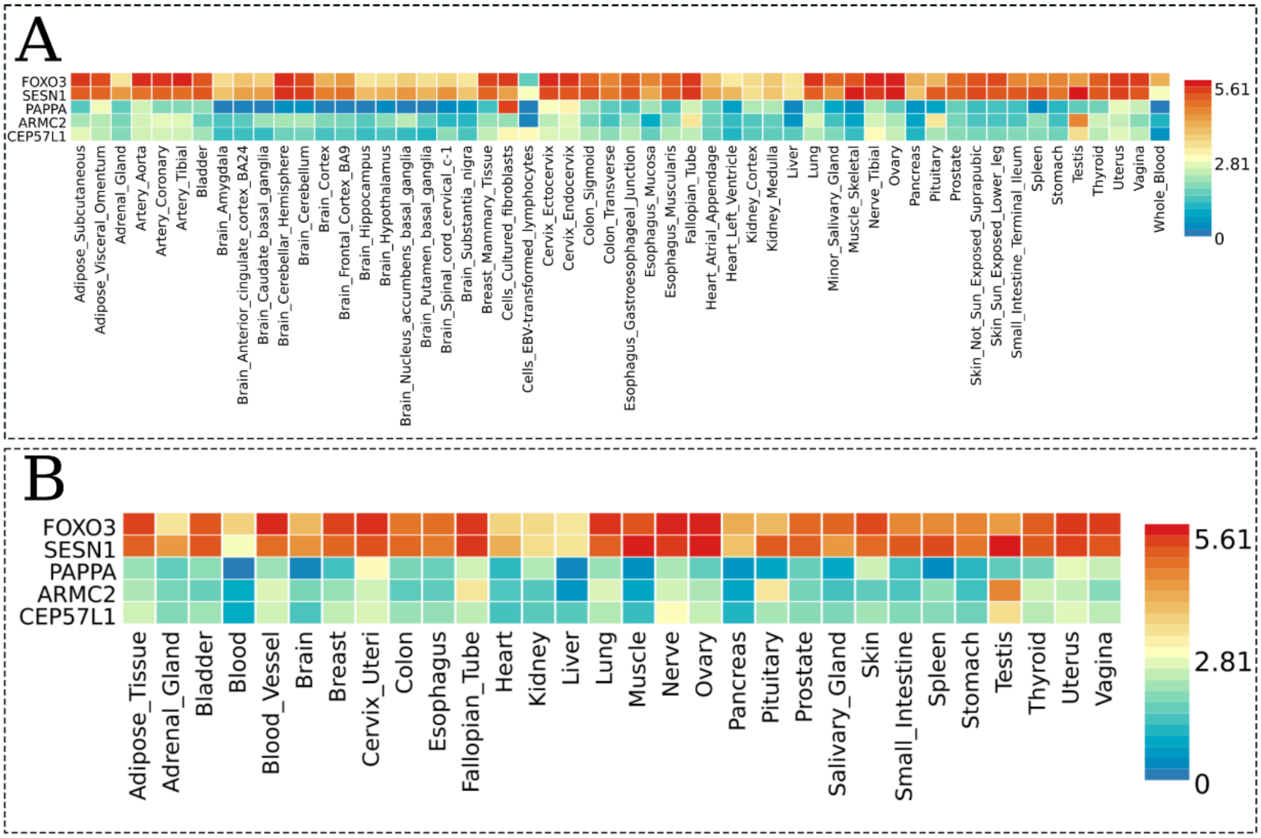


**Fig. S10: Results from tissue-specific gene expression analyses for the A2 dimension.** Heatmaps display the average expression value ($log_2$ transformed) for the prioritized genes (determined by physical position mapping of the GWAS lead SNPs) in the GTEx v8 RNAseq data: (A) 54 tissue types and (B) 30 general tissue types. Red cells indicate relatively high expressions of the corresponding genes in the tissue compared to the blue cells. The prioritized genes were not included in the selected expression data for the A1 and A3 dimensions.

**Table S8.** Site breakdown of the ABIDE consortium analyzed.

| No | Site | From | Total | ASD (M/F) | TD† (M/F) |
|---|---|---|---|---|---|
| 1 | Barrow Neurological Institute | ABIDE 2 | 57 | 28 (28/0) | 29 (29/0) |
| 2 | California Institute of Technology | ABIDE 1 | 37 | 19 (15/4) | 18 (14/4) |
| 3 | Carnegie Mellon University | ABIDE 1 | 27 | 14 (11/3) | 13 (10/3) |
| 4 | ETH Zurich | ABIDE 2 | 34 | 11 (11/0) | 23 (23/0) |
| 5 | Indiana University | ABIDE 2 | 36 | 20 (16/4) | 16 (15/1) |
| 6 | Institut Pasteur and Robert Debre Hospital | ABIDE 2 | 32 | 8 (4/4) | 24 (9/15) |
| 7 | Institute of Living at Hartford Hospital | ABIDE 2 | 49 | 23 (19/4) | 26 (20/6) |
| 8 | Ludwig Maximilians University Munich | ABIDE 1 | 43 | 16 (13/3) | 27 (23/4) |
| 9 | Netherlands Institute for Neurosciences | ABIDE 1 | 30 | 15 (15/0) | 15 (15/0) |
| 10 | New York University | ABIDE 1&2 | 66 | 25 (20/5) | 41 (33/8) |
| 11 | San Diego State University | ABIDE 1&2 | 25 | 13 (11/2) | 12 (11/1) |
| 12 | Trinity Centre for Health Sciences | ABIDE 1 | 25 | 13 (13/0) | 12 (12/0) |
| 13 | University of Leuven | ABIDE 1 | 34 | 15 (15/0) | 19 (19/0) |
| 14 | University of Michigan | ABIDE 1 | 38 | 10 (7/3) | 28 (21/7) |
| 15 | University of Pittsburgh School of Medicine | ABIDE 1 | 32 | 18 (18/0) | 14 (13/1) |
| 16 | University of Utah School of Medicine | ABIDE 1&2 | 104 | 59 (57/2) | 45 (42/3) |
| | **Total** | | 669 | 307 (273/34) | 362 (309/53) |

*M: males; F: females

†Only sites with the number of typically developing (TD) controls greater than 10 within the age range (16-64 years) were included in the analyses.

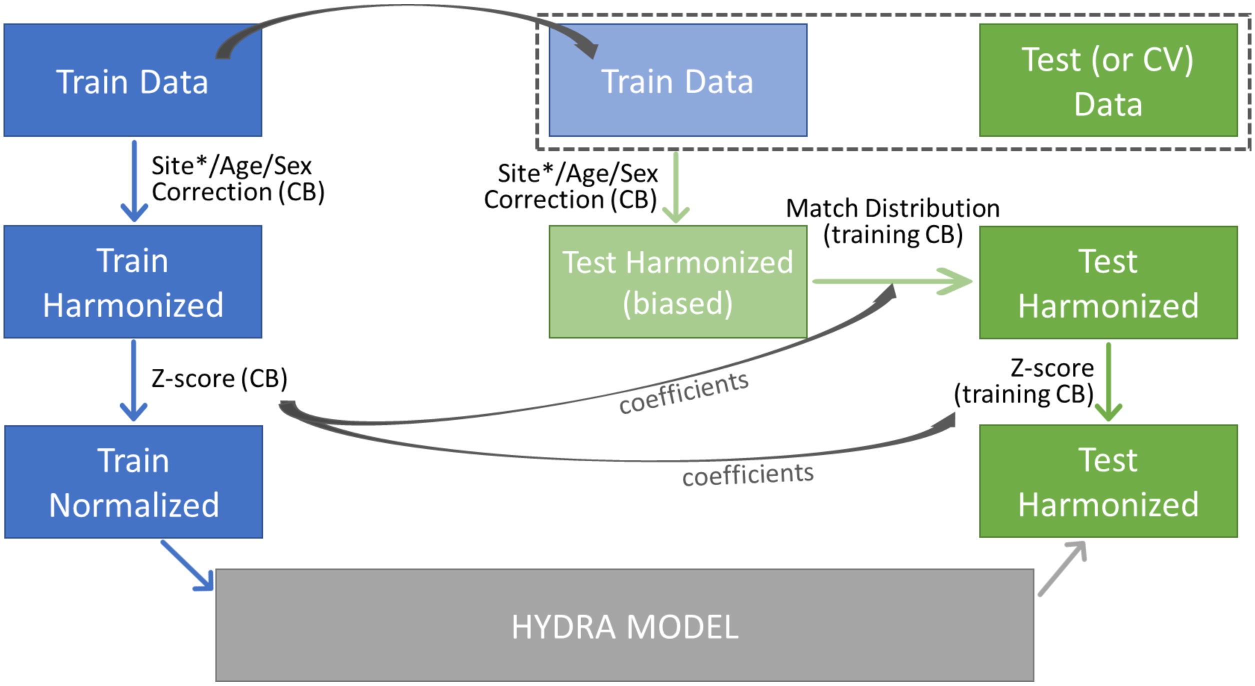


**Fig. S11: Flowchart of the image harmonization.** This flowchart summarizes the image harmonization process for the training datasets (left, blue) and also for the testing or cross validation (CV) datasets (right, green). The harmonization for testing datasets was performed with including the corresponding training dataset, and then the distributions of the harmonized ROIs of the control population in the training dataset were matched, to remove any bias such as those caused by differing age ranges or sex ratios between the training and testing datasets. (CB: control-based) *Site harmonization (ComBat) was run separately, prior to age/sex correction, with age, sex, and the intracranial volume (ICV) as covariates[14,15].

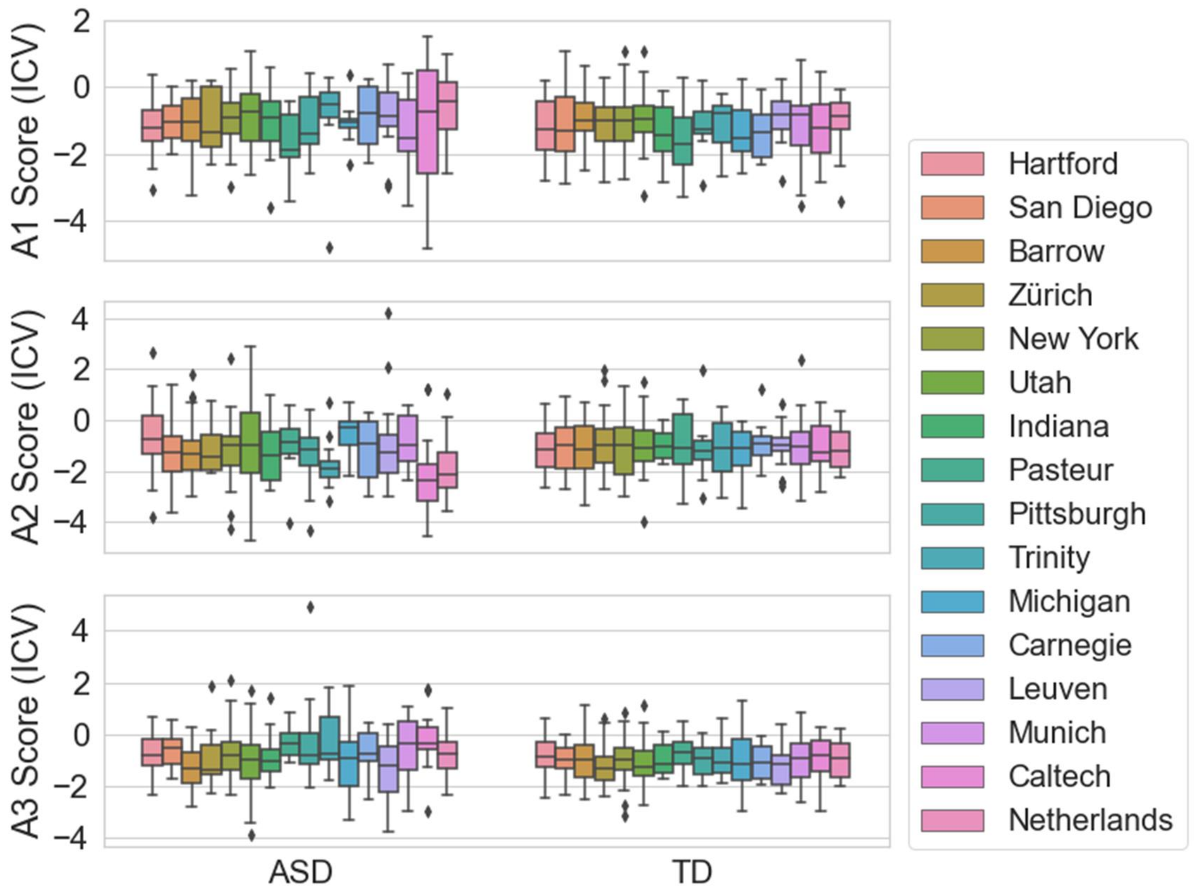


**Fig. S12: ICV-corrected dimension scores by site and diagnosis.** No statistical differences were found in the three ASD dimension scores of TD controls between 16 sites, after the scores were adjusted for the intracranial volume (ICV) ($P$>0.9, one-way ANOVA). This confirms the effectiveness of control-based ComBat site harmonization. Significant differences were found in the A2 scores of individuals with ASD ($P$=0.19), while no sigficance in the other two ($P$>0.1)
