## Supplementary figures and images for "Three Imaging Endophenotypes Characterize Neuroanatomical Heterogeneity of Autism Spectrum Disorder"

### Supplementary Video 1

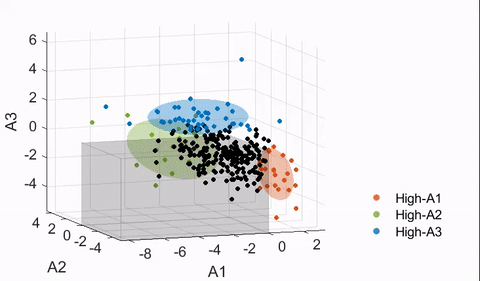
